# Cross-sectional and longitudinal associations between sleep disturbances and psycho-affective symptoms in older adults: influence of amyloid positivity

**DOI:** 10.64898/2026.01.15.26344227

**Authors:** Claire André, Edelweiss Touron, Antoine Garnier-Crussard, Andrée-Ann Baril, Gaël Chételat, Sophie Dautricourt, Natalie L. Marchant, Géraldine Rauchs, the Medit-Ageing Research Group

## Abstract

**Background:** Psycho-affective symptoms and repetitive negative thinking (RNT) may increase Alzheimer’s disease (AD) risk. As sleep plays a key role in emotional regulation, we investigated both cross-sectional and longitudinal associations between sleep disturbances and psycho-affective health, according to amyloid-beta (Aβ) status.

**Methods:** One hundred and thirty-four cognitively unimpaired older adults from the Age-Well interventional study (mean age = 68.8 ± 3.8 years; 82 women; 37 Aβ+ individuals) completed psycho-affective (Geriatric Depression Scale, State-Trait Anxiety Inventory), RNT (Rumination Response Scale–brooding, Penn State Worry Questionnaire) and sleep questionnaires (Pittsburgh Sleep Quality Index, Insomnia Severity Index). Wrist actigraphy (n=131; mean recording duration = 7.7 ± 0.5 nights) provided objective measures of sleep fragmentation (sleep fragmentation index, wake time after sleep onset) and their night-to-night variability. Cross-sectional multiple linear regressions and longitudinal linear mixed-effects models (mean follow-up = 3.98 ± 1.15 years) examined associations between sleep and psycho-affective symptoms, adjusting for age, sex, continuous positive airway pressure use, and the non-pharmacological intervention group (for longitudinal analyses).

**Results:** In the whole cohort and in Aβ− individuals, higher self-reported sleep difficulties and insomnia symptoms were cross-sectionally associated with greater anxiety, depression and RNT (all *p*_FDR-corr_<0.05). In Aβ+ individuals, objective sleep fragmentation and instability were cross-sectionally associated with higher anxiety, depression and RNT. Longitudinally, higher baseline insomnia severity, mean sleep fragmentation, and night-to-night sleep instability predicted brooding worsening in Aβ+ individuals only, while poorer perceived sleep quality predicted depressive symptoms worsening (all *p*_FDR-corr_<0.05).

**Conclusions:** In cognitively unimpaired older adults, associations between sleep disturbances and psycho-affective symptoms differ according to Aβ status. In Aβ+ individuals specifically, objective sleep fragmentation and instability are linked to psycho-affective vulnerability and brooding worsening over time. Sleep disturbances could thus represent early modifiable targets to mitigate psycho-affective symptomatology in older adults.

## Introduction

Sleep disturbances are highly prevalent in older adults and increase with age, notably manifesting as higher self-reported sleep difficulties, insomnia symptoms, and objective sleep fragmentation (e.g., increased number of awakenings, increased wake time after sleep onset [WASO] duration) [1,2]. These changes are not only a consequence of the aging process, but are increasingly recognized as markers and potential contributors to neurodegeneration and dementia risk. Converging evidence suggests that alterations in sleep architecture and continuity are detectable even before the onset of neurodegenerative diseases, including Alzheimer’s disease (AD), positioning sleep as a potential early indicator of brain vulnerability [2,3]. Indeed, AD pathology (i.e., amyloid-beta [Aβ] and tau accumulation) emerges decades before the onset of clinical symptoms and accumulates, at the preclinical stage of the disease, in brain regions involved in sleep regulation, such as brainstem nuclei (e.g., the locus coeruleus), basal forebrain and hypothalamus [4–7]. In addition, longitudinal studies also indicate that sleep disturbances may increase AD risk and accelerate the accumulation of AD pathology [8–11], for example through impaired glymphatic clearance [12]. Thus, it is now increasingly acknowledged that the relationship between sleep disturbances and AD pathology is likely bidirectional, with sleep disturbances being both a contributor to, and consequence of, neurodegeneration. However, we still need to better understand which specific aspects of sleep are disrupted in the preclinical stages of the disease, and determine their impact on clinical symptoms. This is crucial for developing sleep interventions aimed at reducing AD risk.

One potential way by which sleep disturbances may increase AD risk is through an interaction with psycho-affective symptoms, which are themselves recognized as risk factors for dementia [13,14]. First, anxiety and depression are the most prevalent neuropsychiatric manifestations in older adults [15–18]. Anxious and depressive symptoms (even at mild and/or subclinical levels) not only impair quality of life, but some studies also report significant associations with brain changes suggestive of AD in cognitively unimpaired older adults [19–23]. Second, repetitive negative thinking (RNT), a transdiagnostic cognitive process encompassing brooding (i.e., past-directed negative thoughts) and worry (i.e., future-directed negative thoughts), is also emerging as a risk factor for cognitive decline. Indeed, RNT has been linked to neurodegeneration and AD pathology in older adults, as well as cognitive decline [24–27]. Still, the mechanisms and factors influencing both the severity and worsening of psycho-affective symptoms associated with dementia risk need to be further clarified.

Importantly, sleep disturbances may act as a mechanistic bridge between emotional dysregulation and neurodegeneration. It is well-established that sleep disturbances are prominent in psychiatric disorders [28], such as major depressive disorder [28–31]. Moreover, studies conducted in large cohorts of older adults have reported that sleep disturbances are associated with the worsening of anxiety and depressive symptoms over time [32–35]. Emerging evidence also suggests an association between RNT and poor sleep. A previous study from our team showed that higher levels of RNT were associated with lower sleep efficiency measured using polysomnography in cognitively unimpaired older adults [36]. Together, these converging lines of evidence point towards an intersection between sleep, psycho-affective symptoms and AD. Still, most previous research has studied the relationships between sleep, psycho-affective symptoms, and Aβ pathology independently, leaving open the question of whether the interplay between sleep and emotional dysregulation differs as a function of progressing AD pathology. Specifically, whether early cross-sectional associations between sleep disturbances and psycho-affective symptoms differ according to Aβ status in older adults without dementia needs to be established. Furthermore, longitudinal studies offering insights into the potential direction of these associations in the presence or absence of AD pathology are lacking. Addressing these gaps is crucial to delineate early pathways through which sleep disturbances may reflect and/or contribute to behavioural symptoms and increased AD risk, and to help identify early targets for intervention.

The present study aimed to investigate cross-sectional and longitudinal associations between sleep disturbances – including self-reported sleep difficulties, insomnia symptoms, indices of objective sleep fragmentation and their night-to-night variability – and psycho-affective symptoms (i.e., anxiety and depressive symptoms) and RNT in cognitively unimpaired older adults. Importantly, we examined whether these relationships differ according to baseline Aβ positivity. We hypothesized that poorer sleep quality (i.e., more severe self-reported sleep difficulties, insomnia symptoms, sleep fragmentation, and greater sleep instability) would be associated with higher levels of anxiety, depression, and RNT, and that these associations would be only detectable and/or more pronounced in amyloid-positive (Aβ+) individuals, who may be less resilient to the adverse effects of poor sleep. Moreover, we expected poorer sleep quality at baseline to predict the worsening of psycho-affective symptoms over time, especially in Aβ+ individuals. It is also possible that these relationships may be bidirectional, with more severe psycho-affective symptoms being predictive of the worsening of sleep quality over time, especially in Aβ+ individuals. By addressing these questions, this study seeks to clarify how sleep and mental health interact in older adults, in the context of early AD pathology.

## Material & Methods

### 1. Participants and study design

We used data from the Age-Well randomized controlled trial (RCT) of the Medit-Aging European project, sponsored by the French National Institute of Health and Medical Research (INSERM). The flowchart of the study is presented in **Figure 1**. Between 2016 and 2018, we recruited cognitively unimpaired older adults aged over 65 years living in the community in Caen, France and its surroundings. Participants presented no evidence of major neurologic and psychiatric disorders (notably including substance abuse), unstable chronic diseases, or current medication use interfering with cognition. Of note, individuals with a score at the Montgomery-Åsberg Depression Rating Scale (MADRS) above 6, indicative of mild to major depressive disorder, were excluded from the study.

**Figure 1.**
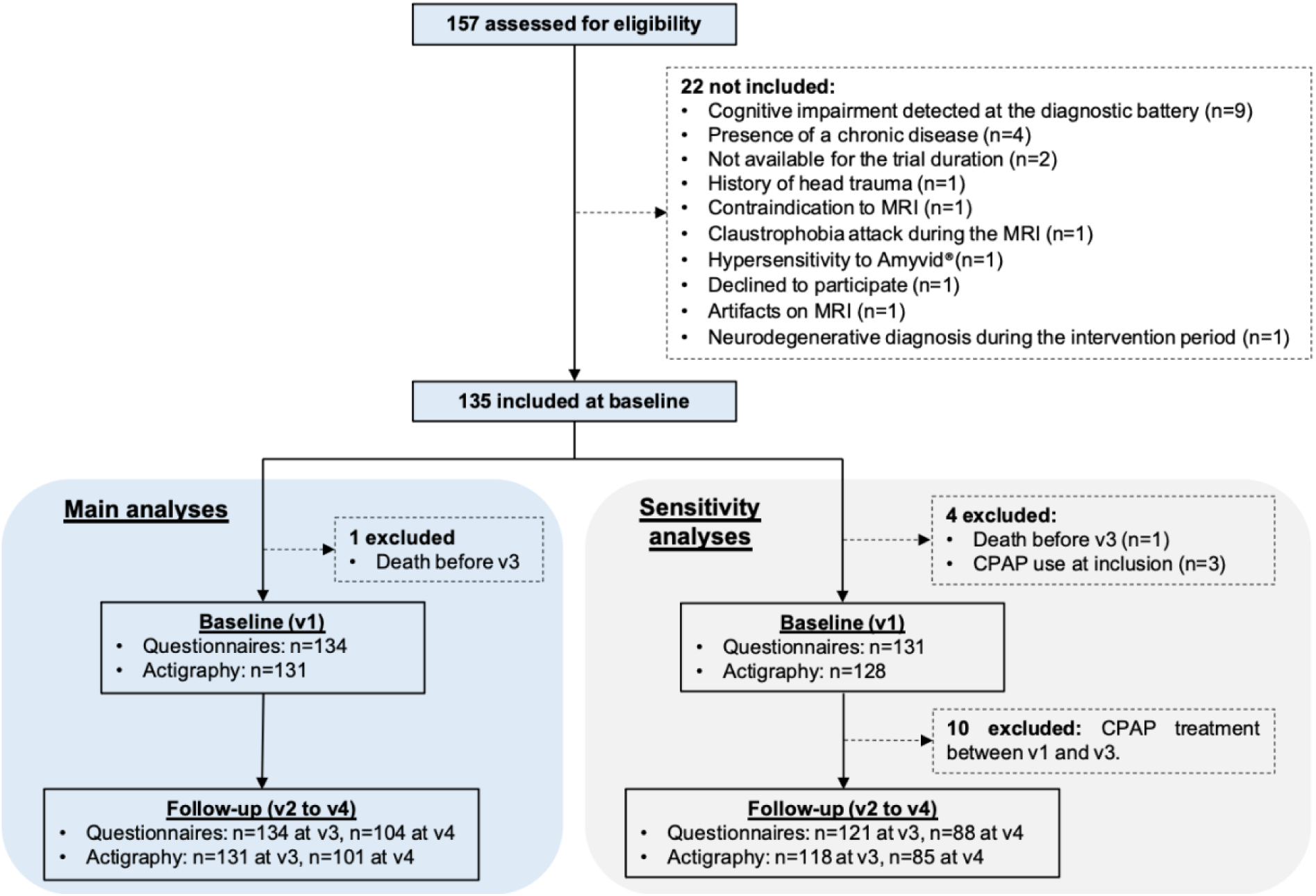
Study flowchart. Flow diagram illustrating participant selection and inclusion, reasons for exclusion, the final main analytic sample, and subsample used for sensitivity analyses. *Abbreviations: CPAP, Continuous Positive Airway Pressure; MRI, Magnetic Resonance Imaging*.

At baseline (visit 1), 135 participants underwent a detailed neuropsychological assessment, structural MRI, ^18^F-Florbetapir PET scan, questionnaires assessing sleep quality and psycho-affective symptoms, and an actigraphy recording within a mean ± SD interval of 2.5 ± 0.9 months (range: 1.1 – 4.4 months). After performing all examinations, they were randomized to one of the three following groups: meditation-based intervention, non-native language training, or passive control arm. Interventions lasted for 18 months. Importantly, as the purpose of the current study is not to study the efficacy of the interventions, we controlled all longitudinal analyses for the intervention group.

A total of 134 participants underwent a longitudinal follow-up with a minimum of 2 post-baseline visits (i.e., visits 2 and 3), and 104 participants underwent an additional visit (i.e., visit 4). The mean ± SD follow-up duration in the cohort is 3.98 ± 1.15 years (range: 1.7 – 4.8 years). At all follow-ups (visits 2, 3 and 4), participants underwent a neuropsychological evaluation and completed psycho-affective questionnaires. Actigraphy recordings were performed again for all participants at visits 3 and 4 (if achieved) only.

All participants included in the Age-Well RCT of the Medit-Aging European project gave their written informed consent before the examinations. The study was granted ethical approval by the “Comité de Protection des Personnes Nord-Ouest III” (Caen, France; trial registration number: EudraCT: 2016-002441-36; IDRCB: 2016-A01767-44; ClinicalTrials.gov Identifier: NCT02977819).

### 2. Sleep measures

#### 2.1. Questionnaires

Self-reported sleep disturbances were assessed using the Pittsburgh Sleep Quality Index (PSQI), a 19-item questionnaire assessing sleep disturbances over the past month [37]. It encompasses 7 components (i.e., sleep quality, sleep onset latency, sleep duration, sleep efficiency, sleep disturbance, use of sleep medication, daytime dysfunction) scored from 0 to 3. A global score ranging from 0 to 21 is calculated as the sum of all components, with higher scores being indicative of greater sleep disturbances. In addition, participants completed the Insomnia Severity Index (ISI), a 7-item questionnaire assessing the severity of current insomnia symptoms [38]. Items cover complementary dimensions of insomnia, including sleep onset difficulties, sleep maintenance difficulties, early morning awakening problems, sleep dissatisfaction, daytime dysfunction, noticeability of sleep problems by others and distress caused by sleep difficulties. Each item is scored from 0 to 4, with the global score ranging from 0 to 28 calculated as the sum of all items, with higher scores being indicative of more severe insomnia symptoms.

#### 2.2. Actigraphy

The sleep-wake cycle was recorded using the MotionWatch 8 wrist-worn triaxial actigraph (CamNTech Ltd, Cambridge, UK), for at least six consecutive nights (range at baseline: 6-8 nights; mean ± SD: 7.7 ± 0.5 nights). As previously described [39], participants were instructed to wear the actigraph on their non-dominant wrist continuously until the end of the recording period. To help with data processing and analysis, they were asked to press the event marker button at lights off and lights on, and to fill in a sleep diary each morning upon awakening [40]. Following the procedure described in [39], data processing was performed using the MotionWare software (version 1.2.14, CamNTech Ltd, Cambridge, UK). Briefly, data were sampled in 5-second epochs, and a sensitivity threshold of 20 counts was applied to distinguish activity from rest. For each recording night, “Lights Off” and “Got Up” markers were positioned by the same experimenter, based on the participant’s event markers and verified using light sensor recordings and sleep diary entries. Subsequently, the “Fell Asleep” and “Woke Up” points were automatically placed by integrating the activity data with the “Lights Off” and “Got Up” markers, respectively, to determine the estimated sleep period. Several indices reflecting sleep fragmentation and night-to-night variability were computed. A mean sleep fragmentation index was calculated by summing the proportions of mobile epochs and immobile bouts lasting ≤1 minute within the identified sleep interval for each night, averaged across all nights for each participant. In addition, we also computed the mean duration of wake time after sleep onset (WASO), as the total length of wake bouts (in minutes) appearing during the sleep period, averaged across all nights for each participant. The night-to-night variability of these parameters was computed as the standard deviation of each parameter across all available recording nights of each individual.

### 3. Assessment of psycho-affective symptoms

Participants completed several scales assessing the presence and severity of psycho-affective symptoms. First, depressive symptoms over the past week were assessed using the 15-item version of the Geriatric Depression Scale (GDS). Scores range from 0 to 15, with higher scores reflecting the presence of more severe depressive symptoms. Second, the State-Trait Anxiety Inventory (STAI) was used to evaluate trait anxiety symptoms, with scores ranging from 20 to 80 and higher scores reflecting more severe anxious symptoms. Third, repetitive negative thinking (RNT) was assessed as a general (trait-like) construct using two scales, reflecting complementary facets of RNT: brooding and worry. Participants completed the 22-item Rumination Response Scale (RRS), and we analysed the 5-item brooding subscale to assess the severity of brooding. Scores range from 5 to 20, with higher scores reflecting greater brooding levels. Worry was evaluated using the 16-item Penn State Worry Questionnaire (PSWQ). Total scores are computed as the sum of the first 11 items and the reverse scores of the latter 5 items, with global scores ranging from 16 to 80 and higher scores reflecting greater worry levels.

### 4. Neuroimaging examinations

A structural MRI and a dual-phase Florbetapir-PET scanning was performed at the Cyceron Center (Caen, France) on a Philips Achieva 3T scanner (Eindhoven, The Netherlands) and a Discovery RX VCT 64 PET-CT scanner (General Electric Healthcare, Milwaukee, WI), respectively. Details on data acquisition and processing have been previously described [41]. Briefly, T1-weighted images were segmented using fluid attenuated inversion recovery (FLAIR) images, spatially normalized to the Montreal Neurologic Institute (MNI) template, modulated using the SPM12 segmentation procedure. PET images were co-registered on corresponding individual T1-weighted MRI scans, spatially normalized to the MNI template using deformation parameters derived from the T1-weighted normalization procedure. Resulting images were quantitatively scaled using cerebellar grey matter as a reference, resulting in standardized uptake value ratio (SUVR) maps. To define Aβ status, average Aβ uptake was extracted from a mask encompassing regions typically exhibiting high Aβ levels in AD (Klunk et al., 2015). Resulting SUVRs were converted in centiloids (CL) following a previously described procedure (Klunk et al., 2015) adapted for Florbetapir SUVRs (Navitsky et al., 2018). Based on previous work, a threshold of 12 CL was used to dichotomize Aβ positive (Aβ+) and negative (Aβ-) individuals (La Joie et al., 2019).

### 5. Statistical analyses

All statistical analyses were conducted using R version 4.5.0 (http://www.r-project.org). The distribution of the variables was visually inspected and checked for normality using Shapiro-Wilk tests. A log transformation was applied to non-normal variables if this improved their distribution and models fit. Differences in demographics, sleep and psycho-affective measures between Aβ+ and Aβ- individuals were assessed using Student t-tests or Mann-Whitney U tests for continuous variables, as appropriate, and chi-squared tests for categorical variables.

#### Cross-sectional analyses

Multiple linear regression analyses were used to assess cross-sectional associations between sleep (predictor) and psycho-affective (outcomes) variables, controlling for age, sex and continuous positive airway pressure (CPAP) treatment for obstructive sleep apnea. These analyses were performed in the whole cohort, as well as in subgroups stratified by Aβ status, separately. All sleep and psycho-affective variables were standardized (Z-scores) to obtain comparable standardized β coefficients across models. Q-Q plots of all regressions were visually inspected to check regression assumptions. To account for multiple testing, p-values were adjusted using the Benjamini–Hochberg false discovery rate (FDR) method. Statistical significance was set to p_FDR-corrected_<0.05.

#### Longitudinal analyses

A first set of longitudinal mixed-effects models assessed the associations between sleep quality at baseline and the evolution of psycho-affective symptoms severity over time, controlling for age, sex, CPAP use and the intervention group. A second set of longitudinal mixed-effects models assessed whether psycho-affective outcomes at baseline predicted longitudinal changes in sleep quality, controlling for the same covariates. All analyses were conducted in the whole cohort and in separate subgroups stratified by Aβ status. P-values were adjusted using an FDR correction, and statistical significance was set to p_FDR-corrected_<0.05.

#### Sensitivity and specificity analyses

As CPAP use may affect both subjective and objective sleep metrics, all cross-sectional analyses were replicated in a subsample from which participants who reported CPAP use at inclusion (n=3) were excluded. Similarly, all longitudinal analyses were replicated after excluding those who started CPAP treatment between v1 and v2 or v3 (n=10). For participants reporting CPAP use at v4, we only analysed v2 and v3 follow-up data.

## Results

### 1. Participants characteristics

One hundred and thirty-four participants were included in the baseline analysis sample, with 131 having valid actigraphy data **(Figure 1)**. Participants’ characteristics and between-group differences (Aβ+ vs Aβ-) at baseline are presented in **Table 1**. Participants had a mean age of 68.82 ± 3.72 years (range: 65-83), a mean level of education of 13.18 ± 3.09 years, 82 (61.2%) were women, and 37 (27.6%) were Aβ+. At baseline, Aβ+ individuals did not differ from Aβ-in terms of demographics, psycho-affective symptoms and cognitive scores, except for marginally lower MMSE scores (U=2171.5, p=0.047). However, Aβ+ participants presented with significantly greater variability in WASO duration and sleep fragmentation index (WASO variability: U=1291.5, p=0.031; Sleep Fragmentation Index variability: T=-2.36, p=0.01). The mean duration of longitudinal follow-up was 3.98 ± 1.15 years, with no between-group difference.

**Table 1.**
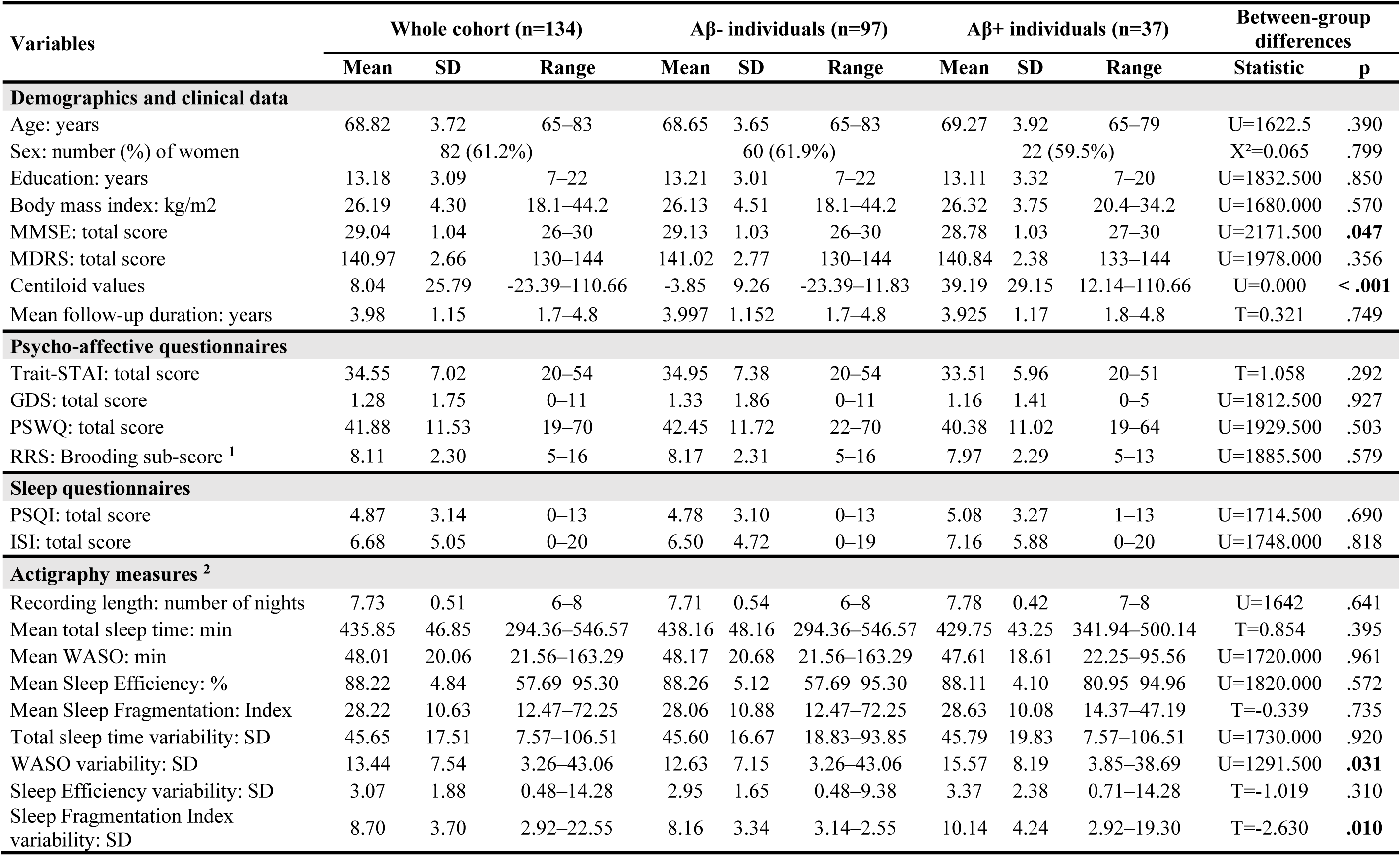
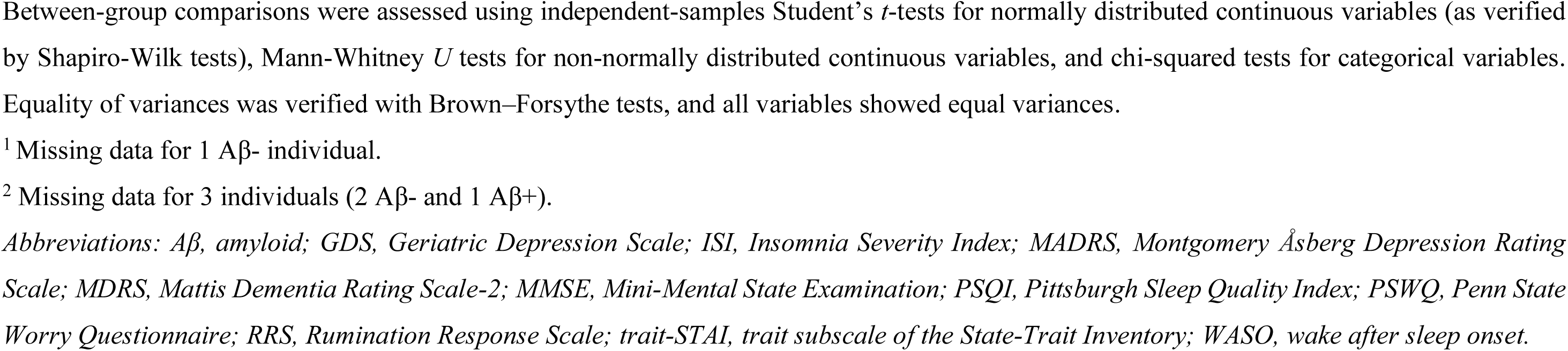
Participants characteristics at baseline.

### 2. Cross-sectional analyses at baseline

#### a. Self-reported sleep measures

In the whole cohort, self-reported sleep disturbances were significantly associated with more severe psycho-affective symptoms. Greater self-reported sleep difficulties, reflected by higher PSQI scores, were significantly associated with more severe anxiety symptoms (Trait-STAI: β=0.25 [95% CI: 0.09 – 0.41], p_FDR-corrected_=0.008) and RNT (RRS brooding: β=0.24 [95% CI: 0.07 – 0.4], p_FDR-corrected_=0.03; PSWQ: β=0.25 [95% CI: 0.08 – 0.41], p_FDR-corrected_=0.01). In addition, higher insomnia symptoms were significantly associated with higher levels of anxiety (Trait-STAI: β=0.41 [95% CI: 0.26 – 0.57], p_FDR-corrected_<0.001), worry (PSWQ: β=0.36 [95% CI: 0.2 – 0.52], p_FDR-corrected_<0.001) and depression (GDS: β=0.24 [95% CI: 0.07 – 0.41], p_FDR-corrected_=0.038) **(Figure 2 and Table S1)**.

**Figure 2.**
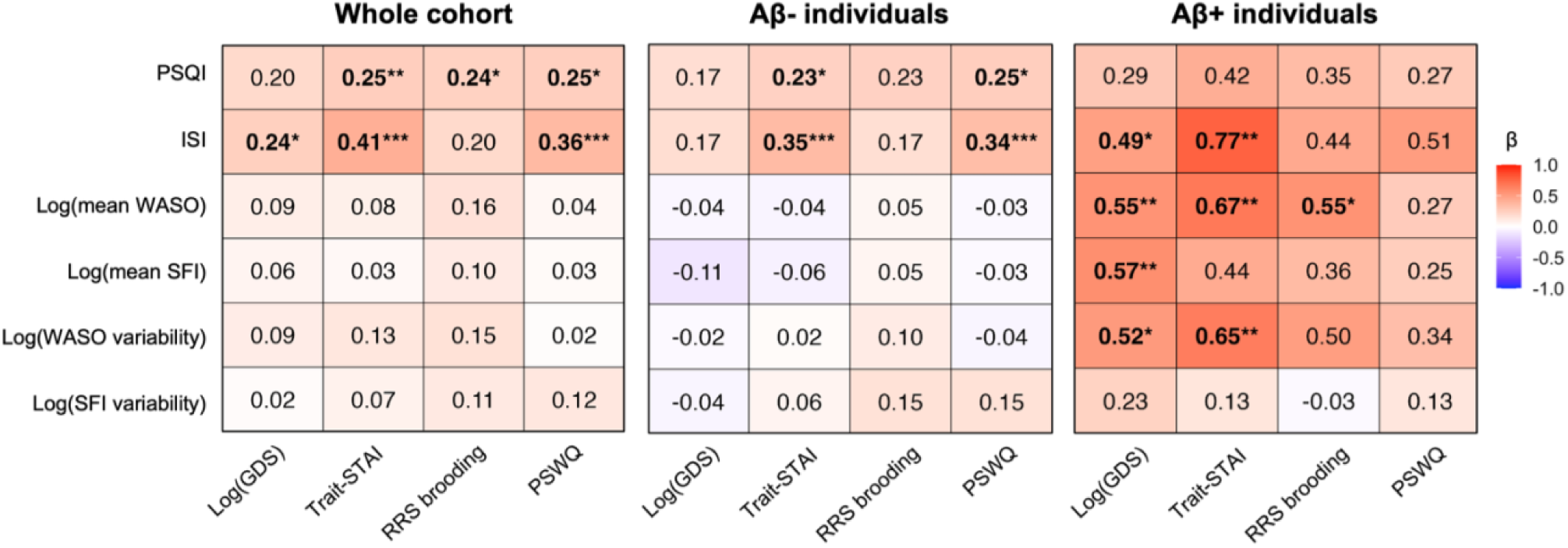
Cross-sectional associations between sleep measures and psycho-affective symptoms. Heatmaps display standardized β coefficients from multiple linear regressions examining associations between self-reported and objective sleep measures (predictors) and psycho-affective symptoms (outcomes), adjusted for age, sex and CPAP use. Analyses were conducted separately in the whole cohort and in subgroups stratified by Aβ status. Results were considered significant when p-values survived an FDR correction for multiple comparisons (*p_FDR-corr_<0.05, **p_FDR-corr_<0.01, ***p_FDR-corr_<0.001). Detailed statistics are available in **Table S1**. *Abbreviations: CPAP, continuous positive airway pressure; FDR, false discovery rate; GDS, Geriatric Depression Scale; ISI, Insomnia Severity Index; PSQI, Pittsburgh Sleep Quality Index; PSWQ, Penn State Worry Questionnaire; RRS, Rumination Response Scale; SD, standard deviation; SFI, sleep fragmentation index; STAI, State-Trait Inventory; WASO, wake after sleep onset*.

When stratifying the sample according to Aβ status, both self-reported sleep difficulties and insomnia symptoms were positively associated with anxiety (PSQI: β=0.23 [95% CI: 0.06 – 0.41], p_FDR-corrected_=0.033; ISI: β=0.35 [95% CI: 0.19 – 0.52], p_FDR-corrected_<0.001) and worry (PSQI: β=0.25 [95% CI: 0.07 – 0.43], p_FDR-corrected_=0.025; ISI: β=0.34 [95% CI: 0.17 – 0.51], p_FDR-corrected_=0.001) in Aβ- individuals **(Figure 2 and Table S1)**. In Aβ+ individuals, insomnia symptoms were positively associated with anxiety and depression levels (Trait-STAI: β=0.77 [95% CI: 0.34 – 1.20], p_FDR-corrected_=0.003; GDS: β=0.49 [95% CI: 0.05 – 0.93], p_FDR-corrected_=0.045) **(Figure 2 and Table S1)**.

#### b. Objective sleep measures

In the whole cohort and in Aβ- individuals, no significant associations were observed between objective sleep metrics and psycho-affective outcomes **(Figure 2 and Table S1)**. However, in Aβ+ individuals, objective sleep disturbances were significantly associated with worse psycho-affective symptoms. In this group only, higher mean wake time after sleep onset (WASO) duration was associated with greater levels of anxiety (β=0.67 [95% CI: 0.32 – 1.02], p_FDR-corrected_=0.003), brooding (β=0.55 [95% CI: 0.19 – 0.92], p_FDR-corrected_=0.025) and depression (β=0.55 [95% CI: 0.21 – 0.89], p_FDR-corrected_=0.007) **(Figure 2 and Table S1)**. In addition, higher mean sleep fragmentation was associated with greater depressive symptoms (β=0.57 [95% CI: 0.22 – 0.93], p_FDR-corrected_=0.007). Finally, higher night-to-night variability of wake time after sleep onset was associated with significantly higher anxiety levels (β=0.65 [95% CI: 0.26 – 1.04], p_FDR-corrected_=0.004) and depressive symptoms (β=0.52 [95% CI: 0.14 – 0.90], p_FDR-corrected_=0.018) **(Figure 2 and Table S1)**.

### 3. Longitudinal analyses

#### a. Baseline sleep as a predictor of longitudinal psycho-affective changes

No longitudinal mixed-effects models were significant after FDR correction for multiple comparisons in the whole cohort or Aβ- individuals **(Table S2)**.

However, in Aβ+ participants only, higher baseline insomnia severity (Baseline ISI: β=0.04, p_FDR-corrected_=0.036), mean sleep fragmentation index (Baseline mean Sleep Fragmentation Index: β=2.11, p_FDR-corrected_=0.012), night-to-night variability in wake time after sleep onset (Baseline WASO variability: β=1.62, p_FDR-corrected_=0.012) and night-to-night variability of sleep fragmentation (Baseline Sleep Fragmentation Index variability: β=1.74, p_FDR-corrected_=0.012) were all significantly associated with the worsening of brooding over time **(Figure 3 and Table S2)**. In addition, higher self-reported sleep difficulties at baseline were significantly associated with a worsening of depressive symptoms (Baseline PSQI: β=0.01, p_FDR-corrected_=0.048).

**Figure 3.**
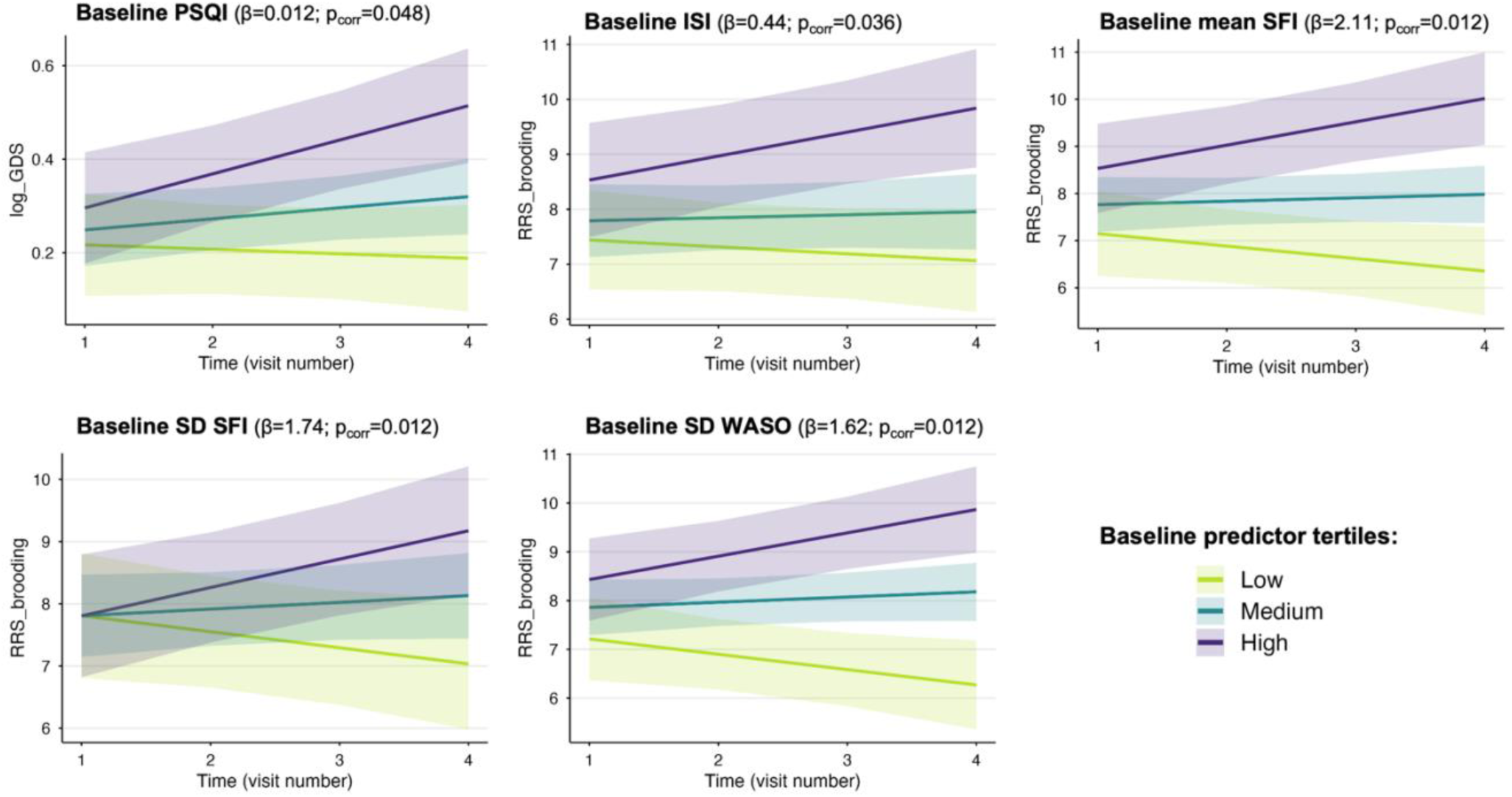
Predicted trajectories from longitudinal mixed-effects models in Aβ+ individuals. Results of significant longitudinal mixed-effects models testing the associations between baseline sleep variables and longitudinal changes in psycho-affective scores in Aβ+ individuals. Predicted values represent model-based marginal estimates (± 95% confidence interval) adjusted for age, sex, CPAP use and the intervention group. Predictions are shown for the mean values of the lower, middle and upper tertiles of each baseline sleep measure. Full statistical details are provided in **Table S2**. *Abbreviations: CPAP, continuous positive airway pressure; GDS, Geriatric Depression Scale; ISI, Insomnia Severity Index; PSQI, Pittsburgh Sleep Quality Index; RRS, Rumination Response Scale; SD, standard deviation; SFI, sleep fragmentation index; WASO, wake after sleep onset*.

#### b. Baseline psycho-affective symptoms as predictors of longitudinal sleep changes

No baseline psycho-affective score was significantly associated with longitudinal changes in sleep outcomes over time after FDR correction in the whole cohort, nor in subgroups stratified by Aβ status **(Table S3)**.

### 4. Sensitivity and specificity analyses

All analyses were replicated in a subsample from which CPAP users were excluded. All cross-sectional results remained unchanged **(Figure S1, Table S4)**. Regarding longitudinal analyses **(Tables S5 and S6)**, results remained unchanged in the whole cohort and in Aβ- participants. However, only two significant associations remained significant in Aβ+ participants: (i) higher self-reported sleep difficulties (reflected by higher PSQI scores) at baseline were still significantly associated with a worsening of depressive symptoms (β=0.01, p_FDR-corrected_=0.032), and (ii) higher sleep fragmentation variability was still significantly associated with the worsening of brooding over time (β=1.71, p_FDR-corrected_=0.049) **(Figure S2, Table S5)**. Of note, the associations between higher baseline insomnia symptoms, and greater baseline wake after sleep onset variability and the worsening of brooding over time became trends (Baseline ISI: β=0.04, p_unc._=0.035, p_FDR-corrected_=0.071; Baseline WASO variability: β=1.25, p_unc.=_0.020, p_FDR-corrected_=0.059) **(Table S5)**.

## Discussion

The goal of the present study was to investigate how sleep disturbances relate to psycho-affective symptoms in cognitively unimpaired older adults, and how these relationships differ according to Aβ status. In the whole sample and in Aβ- participants, higher self-reported sleep difficulties and insomnia symptoms were mainly associated with higher anxiety and RNT at baseline, but no longitudinal associations were observed. In Aβ+ individuals, higher insomnia symptoms were also associated with greater anxiety and depressive symptoms at baseline. Moreover, objective markers of sleep fragmentation and instability were associated with psycho-affective symptoms in Aβ+ individuals only. Specifically, higher mean sleep fragmentation and/or wake time after sleep onset were cross-sectionally associated with higher levels of anxiety, depression, and RNT, while increased night-to-night variability in sleep continuity was associated with higher anxiety and depressive symptoms. When looking at longitudinal data, greater mean sleep fragmentation, instability and insomnia symptoms at baseline predicted worsening of brooding over time, and higher self-reported sleep difficulties predicted worsening of depressive symptoms over time in Aβ+ individuals.

The fact that self-reported sleep disturbances and insomnia symptoms were related to greater anxiety, depression and RNT levels at baseline aligns with previous work highlighting such associations between perceived poor sleep and affective vulnerability. In older adults, significant associations between more severe anxiety symptoms and both higher self-reported sleep difficulties (as reflected by higher PSQI scores) and excessive daytime sleepiness have been previously reported [42]. Moreover, insomnia prevalence is known to increase with advancing age [43], and to be highly comorbid with anxiety in older adults [44,45]. These associations may arise from cognitive–emotional hyperarousal, and involve dysregulation within the prefrontal–limbic circuitry and default mode network, notably the medial prefrontal cortex, anterior cingulate cortex, and amygdala, which support emotion regulation [46]. Besides, a previous meta-analysis has reported that higher RNT (especially ruminations) was associated with worse self-reported sleep quality in young and middle-aged adults, with a medium effect size [47]. Heightened RNT may reflect inefficient top-down control from prefrontal areas over limbic activity, fostering sustained negative affect and physiological arousal that compromise sleep quality. Interestingly, this meta-analysis also reported that RNT was more closely related to self-reported sleep quality rather than actigraphy-derived metrics [47]. Of note, cross-sectional associations between self-reported sleep and psycho-affective disturbances were relatively attenuated in Aβ+ individuals, as only insomnia symptoms were significantly associated with greater anxiety and depressive symptoms in this group.

However, actigraphy-derived indices of sleep fragmentation and instability and psycho-affective symptoms were significantly associated with more severe psycho-affective symptoms in Aβ+ individuals. In this group, higher mean sleep fragmentation indices were associated with higher RNT, anxiety and depressive symptoms, while higher night-to-night sleep variability was associated with higher anxiety and depressive symptoms. In addition, Aβ+ participants also presented with higher night-to-night sleep variability compared to Aβ- participants, which is consistent with several other studies showing associations with greater levels of AD pathology (e.g., higher PET-measured amyloid burden, higher CSF p-tau181/Aβ_42_, higher plasma p-tau231/Aβ_42_) [48–50], and lower global cognitive performance and inhibitory control [49]. Together, our findings support the emerging body of work suggesting that night-to-night sleep variability may be altered in the context of early amyloid deposition, and provide significant new insights by showing its association with greater psycho-affective symptoms. The absence of associations between objective sleep indices and psycho-affective symptoms in Aβ-individuals may be explained by the fact that more subtle alterations in sleep microstructure, that cannot be captured using actigraphy, may be associated with psycho-affective disturbances in this group. For example, rapid eye movement (REM) sleep is known to be intimately involved in emotion regulation [51], such that alterations of REM sleep microstructure could be more closely associated with psycho-affective symptoms in cognitively healthy Aβ-individuals.

Furthermore, longitudinal analyses in Aβ+ individuals show that higher insomnia symptoms, mean sleep fragmentation and sleep instability at baseline were all associated with brooding worsening over time. These longitudinal findings suggest that, in the presence of amyloid pathology, disturbances in sleep continuity may play a role in the worsening of maladaptive RNT. Importantly, the absence of reverse association, whereby baseline psycho-affective symptoms did not predict subsequent sleep changes, further supports the idea that sleep has a specific influence on the emergence and/or worsening of psycho-affective symptoms, in the context of early AD-related pathological changes. At the neurobiological level, it is possible that early AD pathology, especially tau accumulation, in brain regions involved in sleep-wake regulation (e.g., the hypothalamus, thalamus, brainstem nuclei) may destabilize the sleep-wake cycle and underlie greater sleep fragmentation and night-to-night variability in sleep patterns [5–7]. In turn, sleep disturbances could impair neural circuits supporting emotional regulation, cognitive control, and stress responsivity, becoming a driver of psycho-affective symptomatology in preclinical AD. In addition, our findings also show that higher self-perceived sleep difficulties (as measured by greater PSQI scores) at baseline were predictive of depressive symptoms worsening in Aβ+ individuals. In this context, self-reported sleep difficulties may reflect heightened sleep-related hyperarousal, negative cognitive bias, or altered interoceptive processing, and future studies should investigate other objective sleep dimensions to clarify these associations (e.g., alterations in circadian rhythms, reduced slow-wave sleep). Overall, the fact that worse subjective and objective sleep quality at baseline was predictive of the worsening of brooding and depressive symptoms over time only in Aβ+ individuals may have important clinical implications. This suggests that in individuals at greater risk of AD, interventions aimed at reducing insomnia symptoms and stabilizing the sleep-wake rhythm and sleep architecture (e.g., Cognitive Behavioural Therapy for Insomnia, chronobiological interventions such as bright light therapy, pharmacological modulation of sleep continuity for example with Dual Orexin Receptor Antagonists [DORAs] or melatonin, non-invasive neuromodulatory approaches) may help regulate emotional functioning and mitigate psycho-affective decline [52,53].

The present study has important strengths, including the use of both questionnaires and actigraphy to assess complementary aspects of sleep health, a longitudinal design over a mean duration of 4 years, and the stratification of participants according to amyloid status. However, some limitations should also be acknowledged. As participants were cognitively unimpaired, the Aβ+ group formed a limited sample, which may have reduced statistical power for detecting subtle effects. Furthermore, actigraphy, while ecologically valid and suited to assess night-to-night sleep variability, cannot differentiate sleep stages or capture microstructural sleep features, which may be more closely related to psycho-affective health. In this context, replicating current findings using EEG-based wearable devices will represent an important next step. Lastly, the severity of psycho-affective disturbances in our sample was largely sub-clinical overall, with low variability for some measures such as the GDS measuring depressive symptoms. Whether sleep disturbances predict the onset of more severe neuropsychiatric symptoms in later stages of AD and related dementias remains to be clarified in future studies.

## Conclusion

Overall, our findings indicate that the relationship between sleep disturbances and psycho-affective symptoms differs according to amyloid status in cognitively unimpaired older adults. While self-perceived sleep difficulties were primarily associated with anxiety and RNT in Aβ− individuals, Aβ+ individuals showed a specific vulnerability to objective sleep fragmentation and instability. In this group, sleep fragmentation and instability were cross-sectionally associated with psycho-affective symptoms, and longitudinally predicted worsening brooding, suggesting that sleep disturbances may represent an early and modifiable driver of psycho-affective vulnerability in individuals at increased risk of AD.

## Acknowledgements

The authors thank all participants of the study, as well as Stéphane Réhel, Pierre Champetier and Valentin Ourry for their help with sleep data acquisition, Géraldine Poisnel for her administrative support, the Cyceron neuroimaging staff, Euclid team, and the sponsor (INSERM) for their help with data acquisition.

## Data availability statement

Data are made available on request following a formal data sharing agreement and approval by the consortium and executive committee (silversantestudy.eu/2020/09/25/data-sharing). The material can be mobilized, under the conditions and modalities defined in the Medit-Aging Charter, by any research team belonging to an Academic for conducting a scientific research project relating to the scientific theme of mental health and well-being in older people. The material may also be mobilized by non-academic third parties, under conditions, particularly financial, which will be established by a separate agreement between INSERM and by the said third party. Data sharing policies described in the Medit-Aging Charter are in compliance with our ethical approval and guidelines from our funding body.

