## Supplementary Material for "Cross-sectional and longitudinal associations between sleep disturbances and psycho-affective symptoms in older adults: influence of amyloid positivity"

**Table S1**. **Cross-sectional associations between sleep measures and psycho-affective symptoms at baseline.**

**Table S2**. **Associations between baseline sleep measures and longitudinal psycho-affective changes.**

**Table S3**. **Associations between baseline psycho-affective symptoms and sleep changes over time.**

**Figure S1**. **Heatmaps of cross-sectional associations between sleep measures and psycho-affective symptoms after exclusion of CPAP users at baseline.**

**Figure S2**. **Predicted trajectories from significant longitudinal mixed-effects models in Aβ+ individuals, after exclusion of CPAP users.**

**Table S4**. **Cross-sectional associations between sleep measures and psycho-affective symptoms at baseline, after exclusion of CPAP users.**

**Table S5**. **Associations between baseline sleep measures and longitudinal psycho-affective changes, after exclusion of CPAP users.**

**Table S6**. **Associations between baseline psycho-affective symptoms and sleep changes over time, after exclusion of CPAP users.**

**Appendix**. **The Medit-Ageing Research Group.**

**Table S1. Cross-sectional associations between sleep measures and psycho-affective symptoms at baseline.**

| Dependent variable | Independent variable | Adjusted $R^2$ | Beta | 95% CI | SE | t | $P_{unc.}$ | $P_{FDRcorr}$ |
|---|---|---|---|---|---|---|---|---|
| **WHOLE COHORT (n=134)** | | | | | | | | |
| ISI | Log_GDS | 0.08 | 0.24 | 0.07 – 0.42 | 0.09 | 2.77 | 0.006 | **0.038** |
| ISI | Trait-STAI | 0.20 | 0.41 | 0.26 – 0.57 | 0.08 | 5.28 | <0.001 | **<0.001** |
| ISI | RRS brooding | 0.06 | 0.20 | 0.03 – 0.36 | 0.08 | 2.32 | 0.022 | 0.066 |
| ISI | PSWQ | 0.16 | 0.36 | 0.20 – 0.52 | 0.08 | 4.49 | <0.001 | **<0.001** |
| PSQI | Log_GDS | 0.09 | 0.20 | 0.02 – 0.37 | 0.09 | 2.25 | 0.026 | 0.078 |
| PSQI | Trait-STAI | 0.12 | 0.25 | 0.09 – 0.41 | 0.08 | 3.05 | 0.003 | **0.008** |
| PSQI | RRS brooding | 0.11 | 0.24 | 0.07 – 0.40 | 0.08 | 2.85 | 0.005 | **0.030** |
| PSQI | PSWQ | 0.12 | 0.25 | 0.08 – 0.41 | 0.08 | 2.96 | 0.004 | **0.011** |
| log_meanWASO | Log_GDS | 0.03 | 0.09 | -0.09 – 0.27 | 0.09 | 0.98 | 0.331 | 0.496 |
| log_meanWASO | Trait-STAI | 0.03 | 0.08 | -0.09 – 0.25 | 0.09 | 0.91 | 0.363 | 0.545 |
| log_meanWASO | RRS brooding | 0.05 | 0.16 | -0.01 – 0.33 | 0.09 | 1.81 | 0.073 | 0.133 |
| log_meanWASO | PSWQ | 0.02 | 0.04 | -0.13 – 0.22 | 0.09 | 0.46 | 0.644 | 0.855 |
| log_meanSFI | Log_GDS | 0.03 | 0.06 | -0.12 – 0.23 | 0.09 | 0.61 | 0.542 | 0.651 |
| log_meanSFI | Trait-STAI | 0.03 | 0.03 | -0.14 – 0.21 | 0.09 | 0.40 | 0.690 | 0.690 |
| log_meanSFI | RRS brooding | 0.03 | 0.10 | -0.07 – 0.28 | 0.09 | 1.20 | 0.233 | 0.233 |
| log_meanSFI | PSWQ | 0.03 | 0.03 | -0.15 – 0.20 | 0.09 | 0.29 | 0.771 | 0.855 |
| log_SD_WASO | Log_GDS | -0.02 | 0.09 | -0.09 – 0.28 | 0.09 | 0.99 | 0.323 | 0.496 |
| log_SD_WASO | Trait-STAI | -0.01 | 0.13 | -0.05 – 0.30 | 0.09 | 1.45 | 0.150 | 0.300 |
| log_SD_WASO | RRS brooding | -0.01 | 0.15 | -0.02 – 0.33 | 0.09 | 1.71 | 0.089 | 0.133 |
| log_SD_WASO | PSWQ | -0.03 | 0.02 | -0.16 – 0.20 | 0.09 | 0.18 | 0.855 | 0.855 |
| log_SD_SFI | Log_GDS | -0.02 | 0.02 | -0.16 – 0.20 | 0.09 | 0.21 | 0.834 | 0.834 |
| log_SD_SFI | Trait-STAI | -0.02 | 0.07 | -0.11 – 0.24 | 0.09 | 0.74 | 0.459 | 0.551 |
| log_SD_SFI | RRS brooding | -0.01 | 0.11 | -0.07 – 0.29 | 0.09 | 1.24 | 0.218 | 0.233 |
| log_SD_SFI | PSWQ | -0.01 | 0.12 | -0.05 – 0.30 | 0.09 | 1.37 | 0.172 | 0.344 |
| **Aβ- INDIVIDUALS (n=97)** | | | | | | | | |
| ISI | Log_GDS | 0.07 | 0.17 | -0.01 – 0.36 | 0.09 | 1.85 | 0.068 | 0.257 |
| ISI | Trait-STAI | 0.20 | 0.35 | 0.19 – 0.52 | 0.08 | 4.32 | <0.001 | **<0.001** |
| ISI | RRS brooding | 0.06 | 0.17 | -0.01 – 0.35 | 0.09 | 1.84 | 0.070 | 0.209 |
| ISI | PSWQ | 0.18 | 0.34 | 0.17 – 0.51 | 0.09 | 4.00 | 0.000 | **0.001** |
| PSQI | Log_GDS | 0.10 | 0.17 | -0.02 – 0.37 | 0.10 | 1.74 | 0.086 | 0.257 |
| PSQI | Trait-STAI | 0.13 | 0.23 | 0.06 – 0.41 | 0.09 | 2.60 | 0.011 | **0.033** |
| PSQI | RRS brooding | 0.12 | 0.23 | 0.05 – 0.42 | 0.09 | 2.47 | 0.015 | 0.092 |
| PSQI | PSWQ | 0.14 | 0.25 | 0.07 – 0.43 | 0.09 | 2.70 | 0.008 | **0.025** |
| log_meanWASO | Log_GDS | 0.00 | -0.04 | -0.25 – 0.17 | 0.11 | -0.36 | 0.716 | 0.815 |
| log_meanWASO | Trait-STAI | 0.00 | -0.04 | -0.24 – 0.15 | 0.10 | -0.45 | 0.651 | 0.781 |
| log_meanWASO | RRS brooding | 0.01 | 0.05 | -0.15 – 0.25 | 0.10 | 0.48 | 0.632 | 0.632 |
| log_meanWASO | PSWQ | 0.00 | -0.03 | -0.23 – 0.18 | 0.10 | -0.28 | 0.783 | 0.792 |
| log_meanSFI | Log_GDS | 0.03 | -0.11 | -0.31 – 0.10 | 0.10 | -1.01 | 0.314 | 0.627 |

| | | | | | | | | |
|---|---|---|---|---|---|---|---|---|
| log_meanSFI | Trait-STAI | 0.02 | -0.06 | -0.25 – 0.14 | 0.10 | -0.59 | 0.559 | 0.781 |
| log_meanSFI | RRS brooding | 0.02 | 0.05 | -0.15 – 0.25 | 0.10 | 0.49 | 0.625 | 0.632 |
| log_meanSFI | PSWQ | 0.02 | -0.03 | -0.23 – 0.17 | 0.10 | -0.26 | 0.792 | 0.792 |
| log_SD_WASO | Log_GDS | -0.04 | -0.02 | -0.23 – 0.18 | 0.10 | -0.24 | 0.815 | 0.815 |
| log_SD_WASO | Trait-STAI | -0.04 | 0.02 | -0.17 – 0.22 | 0.10 | 0.24 | 0.811 | 0.811 |
| log_SD_WASO | RRS brooding | -0.02 | 0.10 | -0.10 – 0.30 | 0.10 | 0.99 | 0.327 | 0.490 |
| log_SD_WASO | PSWQ | -0.04 | -0.04 | -0.24 – 0.16 | 0.10 | -0.42 | 0.675 | 0.792 |
| log_SD_SFI | Log_GDS | -0.04 | -0.04 | -0.24 – 0.16 | 0.10 | -0.44 | 0.663 | 0.815 |
| log_SD_SFI | Trait-STAI | -0.04 | 0.06 | -0.13 – 0.24 | 0.09 | 0.61 | 0.541 | 0.781 |
| log_SD_SFI | RRS brooding | -0.01 | 0.15 | -0.04 – 0.34 | 0.10 | 1.54 | 0.128 | 0.256 |
| log_SD_SFI | PSWQ | -0.01 | 0.15 | -0.04 – 0.34 | 0.10 | 1.57 | 0.121 | 0.242 |
| **Aβ+ INDIVIDUALS (n=37)** | | | | | | | | |
| ISI | Log_GDS | 0.07 | 0.49 | 0.05 – 0.93 | 0.22 | 2.27 | 0.030 | **0.045** |
| ISI | Trait-STAI | 0.24 | 0.77 | 0.34 – 1.20 | 0.21 | 3.67 | 0.001 | **0.003** |
| ISI | RRS brooding | 0.04 | 0.44 | -0.01 – 0.90 | 0.22 | 1.99 | 0.056 | 0.111 |
| ISI | PSWQ | 0.09 | 0.51 | 0.08 – 0.93 | 0.21 | 2.44 | 0.020 | 0.123 |
| PSQI | Log_GDS | -0.01 | 0.29 | -0.12 – 0.70 | 0.20 | 1.43 | 0.162 | 0.194 |
| PSQI | Trait-STAI | 0.04 | 0.42 | -0.01 – 0.85 | 0.21 | 1.97 | 0.057 | 0.068 |
| PSQI | RRS brooding | 0.01 | 0.35 | -0.06 – 0.76 | 0.20 | 1.72 | 0.095 | 0.114 |
| PSQI | PSWQ | -0.02 | 0.27 | -0.13 – 0.67 | 0.20 | 1.38 | 0.176 | 0.268 |
| log_meanWASO | Log_GDS | 0.26 | 0.55 | 0.21 – 0.89 | 0.17 | 3.31 | 0.002 | **0.007** |
| log_meanWASO | Trait-STAI | 0.33 | 0.67 | 0.32 – 1.02 | 0.17 | 3.93 | 0.000 | **0.003** |
| log_meanWASO | RRS brooding | 0.23 | 0.55 | 0.19 – 0.92 | 0.18 | 3.09 | 0.004 | **0.025** |
| log_meanWASO | PSWQ | 0.05 | 0.27 | -0.13 – 0.66 | 0.19 | 1.36 | 0.183 | 0.268 |
| log_meanSFI | Log_GDS | 0.21 | 0.57 | 0.22 – 0.93 | 0.17 | 3.33 | 0.002 | **0.007** |
| log_meanSFI | Trait-STAI | 0.07 | 0.44 | 0.02 – 0.85 | 0.20 | 2.16 | 0.039 | 0.058 |
| log_meanSFI | RRS brooding | 0.03 | 0.36 | -0.06 – 0.77 | 0.20 | 1.77 | 0.087 | 0.114 |
| log_meanSFI | PSWQ | -0.02 | 0.25 | -0.16 – 0.66 | 0.20 | 1.24 | 0.224 | 0.268 |
| log_SD_WASO | Log_GDS | 0.13 | 0.52 | 0.14 – 0.90 | 0.19 | 2.79 | 0.009 | **0.018** |
| log_SD_WASO | Trait-STAI | 0.20 | 0.65 | 0.26 – 1.04 | 0.19 | 3.40 | 0.002 | **0.004** |
| log_SD_WASO | RRS brooding | 0.09 | 0.50 | 0.09 – 0.91 | 0.20 | 2.49 | 0.018 | 0.054 |
| log_SD_WASO | PSWQ | 0.00 | 0.34 | -0.08 – 0.76 | 0.21 | 1.65 | 0.110 | 0.268 |
| log_SD_SFI | Log_GDS | -0.02 | 0.23 | -0.20 – 0.66 | 0.21 | 1.11 | 0.277 | 0.277 |
| log_SD_SFI | Trait-STAI | -0.04 | 0.13 | -0.33 – 0.60 | 0.23 | 0.58 | 0.565 | 0.565 |
| log_SD_SFI | RRS brooding | -0.06 | -0.03 | -0.49 – 0.43 | 0.23 | -0.12 | 0.905 | 0.905 |
| log_SD_SFI | PSWQ | -0.04 | 0.13 | -0.31 – 0.58 | 0.22 | 0.60 | 0.551 | 0.551 |

Results from multiple regression analyses examining cross-sectional associations between self-reported (PSQI, ISI) and objective sleep measures and psycho-affective symptoms at baseline. Data are reported as standardized β coefficients with corresponding 95% confidence intervals (CI), and both uncorrected ($P_{unc.}$) and FDR-corrected ($P_{FDRcorr}$) p-values. Results surviving an FDR correction ($P_{FDRcorr}<0.05$) are indicated in bold. All models were adjusted for age, sex and CPAP use, and analyses were performed in the whole cohort and in subgroups stratified by Aβ status. *Abbreviations: Aβ, amyloid; CPAP, continuous positive airway pressure; FDR, false discovery rate; GDS, Geriatric Depression Scale; ISI, Insomnia Severity Index; PSQI, Pittsburgh Sleep Quality Index; PSWQ, Penn State Worry Questionnaire; RRS, Ruminative Response Scale; SD, standard deviation; SFI, sleep fragmentation index; STAI, State-Trait Anxiety Inventory; WASO, wake after sleep onset.*

**Table S2. Associations between baseline sleep measures and longitudinal psycho-affective changes.**

| Outcome | Time × Predictor interaction | Estimate | SE | t | $P_{unc.}$ | $P_{FDRcorr}$ |
|---|---|---|---|---|---|---|
| **WHOLE COHORT (n=134)** | | | | | | |
| log_GDS | Time x baseline_ISI | 0.001 | 0.00 | 0.76 | 0.45 | 0.61 |
| log_GDS | Time x baseline_PSQI | 0.002 | 0.00 | 0.66 | 0.51 | 0.61 |
| log_GDS | Time x baseline_log_meanWASO | 0.06 | 0.05 | 1.07 | 0.29 | 0.57 |
| log_GDS | Time x baseline_log_meanSFI | 0.10 | 0.06 | 1.82 | 0.07 | 0.41 |
| log_GDS | Time x baseline_log_SD_WASO | 0.01 | 0.04 | 0.35 | 0.73 | 0.73 |
| log_GDS | Time x baseline_log_SD_SFI | 0.07 | 0.05 | 1.35 | 0.18 | 0.53 |
| PSWQ | Time x baseline_ISI | -0.07 | 0.04 | -1.75 | 0.08 | 0.24 |
| PSWQ | Time x baseline_PSQI | -0.11 | 0.06 | -1.75 | 0.08 | 0.24 |
| PSWQ | Time x baseline_log_meanWASO | -0.99 | 1.25 | -0.79 | 0.43 | 0.86 |
| PSWQ | Time x baseline_log_meanSFI | -0.11 | 1.37 | -0.08 | 0.94 | 0.97 |
| PSWQ | Time x baseline_log_SD_WASO | -0.04 | 0.93 | -0.04 | 0.97 | 0.97 |
| PSWQ | Time x baseline_log_SD_SFI | 0.59 | 1.16 | 0.50 | 0.62 | 0.92 |
| RRS brooding | Time x baseline_ISI | 0.02 | 0.01 | 1.81 | 0.07 | 0.14 |
| RRS brooding | Time x baseline_PSQI | 0.01 | 0.02 | 0.62 | 0.54 | 0.54 |
| RRS brooding | Time x baseline_log_meanWASO | 0.40 | 0.39 | 1.05 | 0.30 | 0.36 |
| RRS brooding | Time x baseline_log_meanSFI | 0.91 | 0.42 | 2.18 | 0.030 | 0.14 |
| RRS brooding | Time x baseline_log_SD_WASO | 0.54 | 0.29 | 1.89 | 0.06 | 0.14 |
| RRS brooding | Time x baseline_log_SD_SFI | 0.59 | 0.35 | 1.67 | 0.10 | 0.14 |
| Trait-STAI | Time x baseline_ISI | -0.02 | 0.03 | -0.54 | 0.59 | 0.88 |
| Trait-STAI | Time x baseline_PSQI | 0.01 | 0.05 | 0.28 | 0.78 | 0.92 |
| Trait-STAI | Time x baseline_log_meanWASO | -0.10 | 0.99 | -0.10 | 0.92 | 0.92 |
| Trait-STAI | Time x baseline_log_meanSFI | 1.11 | 1.07 | 1.03 | 0.30 | 0.60 |
| Trait-STAI | Time x baseline_log_SD_WASO | 0.98 | 0.73 | 1.35 | 0.18 | 0.53 |
| Trait-STAI | Time x baseline_log_SD_SFI | 2.37 | 0.90 | 2.62 | 0.009 | 0.055 |
| **Aβ- INDIVIDUALS (n=97)** | | | | | | |
| log_GDS | Time x baseline_ISI | -0.0003 | 0.00 | -0.13 | 0.90 | 0.90 |
| log_GDS | Time x baseline_PSQI | -0.002 | 0.00 | -0.75 | 0.46 | 0.68 |
| log_GDS | Time x baseline_log_meanWASO | 0.07 | 0.06 | 1.17 | 0.24 | 0.68 |
| log_GDS | Time x baseline_log_meanSFI | 0.15 | 0.07 | 2.09 | 0.038 | 0.23 |
| log_GDS | Time x baseline_log_SD_WASO | 0.02 | 0.05 | 0.37 | 0.71 | 0.85 |
| log_GDS | Time x baseline_log_SD_SFI | 0.05 | 0.06 | 0.88 | 0.38 | 0.68 |
| PSWQ | Time x baseline_ISI | -0.08 | 0.05 | -1.65 | 0.10 | 0.30 |
| PSWQ | Time x baseline_PSQI | -0.18 | 0.08 | -2.40 | 0.017 | 0.10 |
| PSWQ | Time x baseline_log_meanWASO | -0.52 | 1.48 | -0.35 | 0.73 | 0.76 |
| PSWQ | Time x baseline_log_meanSFI | -0.61 | 1.63 | -0.37 | 0.71 | 0.76 |
| PSWQ | Time x baseline_log_SD_WASO | 0.35 | 1.12 | 0.31 | 0.76 | 0.76 |
| PSWQ | Time x baseline_log_SD_SFI | 0.65 | 1.44 | 0.45 | 0.65 | 0.76 |

| | | | | | | |
|---|---|---|---|---|---|---|
| RRS brooding | Time x baseline_ISI | 0.01 | 0.02 | 0.49 | 0.63 | 0.98 |
| RRS brooding | Time x baseline_PSQI | -0.01 | 0.02 | -0.33 | 0.74 | 0.98 |
| RRS brooding | Time x baseline_log_meanWASO | 0.11 | 0.46 | 0.24 | 0.81 | 0.98 |
| RRS brooding | Time x baseline_log_meanSFI | 0.39 | 0.50 | 0.77 | 0.44 | 0.98 |
| RRS brooding | Time x baseline_log_SD_WASO | 0.07 | 0.35 | 0.19 | 0.85 | 0.98 |
| RRS brooding | Time x baseline_log_SD_SFI | -0.01 | 0.44 | -0.02 | 0.98 | 0.98 |
| Trait-STAI | Time x baseline_ISI | 0.01 | 0.04 | 0.18 | 0.86 | 0.86 |
| Trait-STAI | Time x baseline_PSQI | 0.03 | 0.06 | 0.60 | 0.55 | 0.82 |
| Trait-STAI | Time x baseline_log_meanWASO | 0.36 | 1.10 | 0.33 | 0.74 | 0.86 |
| Trait-STAI | Time x baseline_log_meanSFI | 0.95 | 1.21 | 0.78 | 0.43 | 0.82 |
| Trait-STAI | Time x baseline_log_SD_WASO | 1.03 | 0.83 | 1.24 | 0.21 | 0.64 |
| Trait-STAI | Time x baseline_log_SD_SFI | 2.44 | 1.06 | 2.31 | 0.022 | 0.13 |
| **Aβ+ INDIVIDUALS (n=37)** | | | | | | |
| log_GDS | Time x baseline_ISI | 0.004 | 0.00 | 1.54 | 0.13 | 0.38 |
| log_GDS | Time x baseline_PSQI | 0.01 | 0.00 | 2.71 | 0.008 | **0.048** |
| log_GDS | Time x baseline_log_meanWASO | 0.01 | 0.09 | 0.10 | 0.92 | 0.99 |
| log_GDS | Time x baseline_log_meanSFI | 0.002 | 0.10 | 0.02 | 0.99 | 0.99 |
| log_GDS | Time x baseline_log_SD_WASO | 0.01 | 0.07 | 0.14 | 0.89 | 0.99 |
| log_GDS | Time x baseline_log_SD_SFI | 0.11 | 0.08 | 1.27 | 0.21 | 0.41 |
| PSWQ | Time x baseline_ISI | -0.05 | 0.07 | -0.78 | 0.44 | 0.84 |
| PSWQ | Time x baseline_PSQI | 0.05 | 0.12 | 0.43 | 0.67 | 0.84 |
| PSWQ | Time x baseline_log_meanWASO | -2.30 | 2.36 | -0.97 | 0.33 | 0.84 |
| PSWQ | Time x baseline_log_meanSFI | 0.97 | 2.51 | 0.39 | 0.70 | 0.84 |
| PSWQ | Time x baseline_log_SD_WASO | -1.38 | 1.78 | -0.78 | 0.44 | 0.84 |
| PSWQ | Time x baseline_log_SD_SFI | 0.09 | 2.14 | 0.04 | 0.97 | 0.97 |
| RRS brooding | Time x baseline_ISI | 0.04 | 0.02 | 2.29 | 0.024 | **0.036** |
| RRS brooding | Time x baseline_PSQI | 0.05 | 0.03 | 1.61 | 0.11 | 0.11 |
| RRS brooding | Time x baseline_log_meanWASO | 1.17 | 0.70 | 1.67 | 0.10 | 0.11 |
| RRS brooding | Time x baseline_log_meanSFI | 2.11 | 0.73 | 2.90 | 0.005 | **0.012** |
| RRS brooding | Time x baseline_log_SD_WASO | 1.62 | 0.51 | 3.18 | 0.002 | **0.012** |
| RRS brooding | Time x baseline_log_SD_SFI | 1.74 | 0.62 | 2.82 | 0.006 | **0.012** |
| Trait-STAI | Time x baseline_ISI | -0.06 | 0.06 | -1.11 | 0.27 | 0.80 |
| Trait-STAI | Time x baseline_PSQI | -0.04 | 0.10 | -0.43 | 0.67 | 0.80 |
| Trait-STAI | Time x baseline_log_meanWASO | -1.31 | 2.11 | -0.62 | 0.54 | 0.80 |
| Trait-STAI | Time x baseline_log_meanSFI | 1.30 | 2.24 | 0.58 | 0.56 | 0.80 |
| Trait-STAI | Time x baseline_log_SD_WASO | 0.29 | 1.59 | 0.18 | 0.86 | 0.86 |
| Trait-STAI | Time x baseline_log_SD_SFI | 1.64 | 1.89 | 0.86 | 0.39 | 0.80 |

Results of longitudinal mixed-effects models testing whether baseline sleep quality predicted longitudinal changes in psycho-affective symptoms over time. Data are reported as estimated standardized β coefficients (estimates), standard error (SE), t-value, and both uncorrected ($P_{unc.}$) and FDR-corrected ($P_{FDRcorr}$) p-values. Results surviving FDR correction ($P_{FDRcorr}<0.05$) are indicated in bold. All models were adjusted for age, sex, CPAP use and the intervention group.

Analyses were performed in the whole cohort and in subgroups stratified by Aβ status. *Abbreviations: Aβ, amyloid; CPAP, continuous positive airway pressure; FDR, false discovery rate; GDS, Geriatric Depression Scale; ISI, Insomnia Severity Index; PSQI, Pittsburgh Sleep Quality Index; PSWQ, Penn State Worry Questionnaire; RRS, Ruminative Response Scale; SD, standard deviation; SFI, sleep fragmentation index; STAI, State-Trait Anxiety Inventory; WASO, wake after sleep onset.*

**Table S3. Associations between baseline psycho-affective symptoms and sleep changes over time.**

| Outcome | Time × Predictor interaction | Estimate | SE | t | $P_{unc.}$ | $P_{FDRcorr}$ |
|---|---|---|---|---|---|---|
| **Whole cohort (n=134)** | | | | | | |
| ISI | Time x baseline_logGDS | 0.568 | 0.46 | 1.23 | 0.22 | 0.59 |
| ISI | Time x baseline_Trait-STAI | 0.005 | 0.02 | 0.29 | 0.77 | 0.77 |
| ISI | Time x baseline_PSWQ | -0.008 | 0.01 | -0.81 | 0.42 | 0.59 |
| ISI | Time x baseline_RRS brooding | 0.040 | 0.05 | 0.77 | 0.44 | 0.59 |
| PSQI | Time x baseline_logGDS | 0.491 | 0.31 | 1.57 | 0.12 | 0.47 |
| PSQI | Time x baseline_Trait-STAI | 0.013 | 0.01 | 1.14 | 0.25 | 0.51 |
| PSQI | Time x baseline_PSWQ | 0.004 | 0.01 | 0.55 | 0.59 | 0.69 |
| PSQI | Time x baseline_RRS brooding | 0.014 | 0.04 | 0.39 | 0.69 | 0.69 |
| log_meanSFI | Time x baseline_logGDS | 0.016 | 0.02 | 1.09 | 0.28 | 0.85 |
| log_meanSFI | Time x baseline_Trait-STAI | 4.37E-04 | 0.001 | 0.80 | 0.42 | 0.85 |
| log_meanSFI | Time x baseline_PSWQ | -8.35E-05 | 3.40E-04 | -0.25 | 0.81 | 0.99 |
| log_meanSFI | Time x baseline_RRS brooding | 2.10E-05 | 0.002 | 0.01 | 0.99 | 0.99 |
| log_meanWASO | Time x baseline_logGDS | 0.023 | 0.02 | 1.51 | 0.13 | 0.49 |
| log_meanWASO | Time x baseline_Trait-STAI | -1.01E-04 | 0.001 | -0.18 | 0.86 | 0.86 |
| log_meanWASO | Time x baseline_PSWQ | -3.13E-04 | 3.45E-04 | -0.91 | 0.37 | 0.49 |
| log_meanWASO | Time x baseline_RRS brooding | -0.002 | 0.002 | -0.90 | 0.37 | 0.49 |
| log_SD_SFI | Time x baseline_logGDS | 0.011 | 0.02 | 0.46 | 0.65 | 0.65 |
| log_SD_SFI | Time x baseline_Trait-STAI | 0.001 | 0.001 | 1.53 | 0.13 | 0.25 |
| log_SD_SFI | Time x baseline_PSWQ | 2.79E-04 | 0.001 | 0.51 | 0.61 | 0.65 |
| log_SD_SFI | Time x baseline_RRS brooding | 0.005 | 0.003 | 1.77 | 0.08 | 0.25 |
| log_SD_WASO | Time x baseline_logGDS | 0.005 | 0.03 | 0.19 | 0.85 | 0.85 |
| log_SD_WASO | Time x baseline_Trait-STAI | -0.001 | 0.001 | -1.16 | 0.25 | 0.49 |
| log_SD_WASO | Time x baseline_PSWQ | -3.81E-04 | 0.001 | -0.61 | 0.54 | 0.72 |
| log_SD_WASO | Time x baseline_RRS brooding | -0.004 | 0.003 | -1.23 | 0.22 | 0.49 |
| **Aβ- INDIVIDUALS (n=97)** | | | | | | |
| ISI | Time x baseline_logGDS | 0.542 | 0.558 | 0.97 | 0.33 | 0.67 |
| ISI | Time x baseline_Trait-STAI | 0.008 | 0.020 | 0.39 | 0.70 | 0.70 |
| ISI | Time x baseline_PSWQ | -0.016 | 0.012 | -1.27 | 0.21 | 0.67 |
| ISI | Time x baseline_RRS brooding | 0.036 | 0.063 | 0.57 | 0.57 | 0.70 |
| PSQI | Time x baseline_logGDS | 0.356 | 0.346 | 1.03 | 0.30 | 0.62 |
| PSQI | Time x baseline_Trait-STAI | 0.012 | 0.012 | 1.02 | 0.31 | 0.62 |
| PSQI | Time x baseline_PSWQ | -0.002 | 0.008 | -0.29 | 0.77 | 0.85 |
| PSQI | Time x baseline_RRS brooding | 0.007 | 0.039 | 0.19 | 0.85 | 0.85 |
| log_meanSFI | Time x baseline_logGDS | 0.025 | 0.018 | 1.42 | 0.16 | 0.63 |
| log_meanSFI | Time x baseline_Trait-STAI | 0.001 | 0.001 | 0.86 | 0.39 | 0.79 |
| log_meanSFI | Time x baseline_PSWQ | -6.46E-05 | 4.01E-04 | -0.16 | 0.87 | 0.93 |
| log_meanSFI | Time x baseline_RRS brooding | -1.80E-04 | 0.002 | -0.09 | 0.93 | 0.93 |

| | | | | | | |
|---|---|---|---|---|---|---|
| log_meanWASO | Time x baseline_logGDS | 0.027 | 0.017 | 1.58 | 0.12 | 0.47 |
| log_meanWASO | Time x baseline_Trait-STAI | -6.64E-05 | 0.001 | -0.11 | 0.91 | 0.91 |
| log_meanWASO | Time x baseline_PSWQ | -1.45E-04 | 3.84E-04 | -0.38 | 0.71 | 0.91 |
| log_meanWASO | Time x baseline_RRS brooding | -0.001 | 0.002 | -0.77 | 0.44 | 0.88 |
| log_SD_SFI | Time x baseline_logGDS | 0.036 | 0.028 | 1.26 | 0.21 | 0.28 |
| log_SD_SFI | Time x baseline_Trait-STAI | 0.002 | 0.001 | 2.15 | 0.033 | 0.13 |
| log_SD_SFI | Time x baseline_PSWQ | 3.06E-04 | 0.001 | 0.48 | 0.63 | 0.63 |
| log_SD_SFI | Time x baseline_RRS brooding | 0.006 | 0.003 | 1.85 | 0.07 | 0.13 |
| log_SD_WASO | Time x baseline_logGDS | 0.029 | 0.030 | 0.96 | 0.34 | 0.72 |
| log_SD_WASO | Time x baseline_Trait-STAI | -0.001 | 0.001 | -0.61 | 0.54 | 0.72 |
| log_SD_WASO | Time x baseline_PSWQ | -2.23E-04 | 0.001 | -0.33 | 0.74 | 0.74 |
| log_SD_WASO | Time x baseline_RRS brooding | -0.003 | 0.003 | -0.75 | 0.45 | 0.72 |
| **Aβ+ INDIVIDUALS (n=37)** | | | | | | |
| ISI | Time x baseline_logGDS | 0.518 | 0.737 | 0.70 | 0.48 | 0.81 |
| ISI | Time x baseline_Trait-STAI | -0.030 | 0.030 | -1.01 | 0.32 | 0.81 |
| ISI | Time x baseline_PSWQ | 0.004 | 0.016 | 0.24 | 0.81 | 0.81 |
| ISI | Time x baseline_RRS brooding | 0.029 | 0.082 | 0.35 | 0.72 | 0.81 |
| PSQI | Time x baseline_logGDS | 0.823 | 0.666 | 1.24 | 0.22 | 0.69 |
| PSQI | Time x baseline_Trait-STAI | -0.001 | 0.027 | -0.02 | 0.98 | 0.98 |
| PSQI | Time x baseline_PSWQ | 0.014 | 0.015 | 0.95 | 0.34 | 0.69 |
| PSQI | Time x baseline_RRS brooding | 0.017 | 0.074 | 0.23 | 0.82 | 0.98 |
| log_meanSFI | Time x baseline_logGDS | -0.010 | 0.028 | -0.34 | 0.74 | 0.95 |
| log_meanSFI | Time x baseline_Trait-STAI | 6.80E-05 | 0.001 | 0.06 | 0.95 | 0.95 |
| log_meanSFI | Time x baseline_PSWQ | -2.01E-04 | 0.001 | -0.30 | 0.77 | 0.95 |
| log_meanSFI | Time x baseline_RRS brooding | 0.001 | 0.003 | 0.20 | 0.84 | 0.95 |
| log_meanWASO | Time x baseline_logGDS | 0.010 | 0.033 | 0.29 | 0.77 | 0.77 |
| log_meanWASO | Time x baseline_Trait-STAI | -0.001 | 0.001 | -0.37 | 0.71 | 0.77 |
| log_meanWASO | Time x baseline_PSWQ | -0.001 | 0.001 | -1.37 | 0.18 | 0.70 |
| log_meanWASO | Time x baseline_RRS brooding | -0.002 | 0.004 | -0.57 | 0.57 | 0.77 |
| log_SD_SFI | Time x baseline_logGDS | -0.064 | 0.047 | -1.37 | 0.18 | 0.71 |
| log_SD_SFI | Time x baseline_Trait-STAI | -0.002 | 0.002 | -0.93 | 0.35 | 0.71 |
| log_SD_SFI | Time x baseline_PSWQ | -1.03E-04 | 0.001 | -0.09 | 0.93 | 0.93 |
| log_SD_SFI | Time x baseline_RRS brooding | 0.001 | 0.005 | 0.22 | 0.83 | 0.93 |
| log_SD_WASO | Time x baseline_logGDS | -0.075 | 0.062 | -1.21 | 0.23 | 0.26 |
| log_SD_WASO | Time x baseline_Trait-STAI | -0.004 | 0.002 | -1.69 | 0.09 | 0.26 |
| log_SD_WASO | Time x baseline_PSWQ | -0.002 | 0.001 | -1.13 | 0.26 | 0.26 |
| log_SD_WASO | Time x baseline_RRS brooding | -0.010 | 0.007 | -1.36 | 0.18 | 0.26 |

Results of longitudinal mixed-effects models testing whether baseline psycho-affective symptoms predicted longitudinal changes in sleep quality over time. Data are reported as estimated standardized β coefficients (estimates), standard error (SE), t-value, and both uncorrected ($P_{unc.}$) and FDR-corrected ($P_{FDRcorr}$) p-values. Results surviving FDR correction ($P_{FDRcorr}<0.05$) are indicated in bold. All models were adjusted for age, sex, CPAP use and the intervention group. Analyses were performed in the whole cohort and in subgroups stratified by Aβ status. *Abbreviations: Aβ, amyloid; CPAP, continuous positive airway pressure; FDR, false discovery rate; GDS, Geriatric Depression Scale; ISI, Insomnia Severity Index; PSQI, Pittsburgh Sleep Quality Index; PSWQ, Penn State Worry Questionnaire; RRS, Ruminative Response Scale; SD, standard deviation; SFI, sleep fragmentation index; STAI, State-Trait Anxiety Inventory; WASO, wake after sleep onset.*

**<u>Figure S1</u>. Heatmaps of cross-sectional associations between sleep measures and psycho-affective symptoms after exclusion of CPAP users at baseline.**

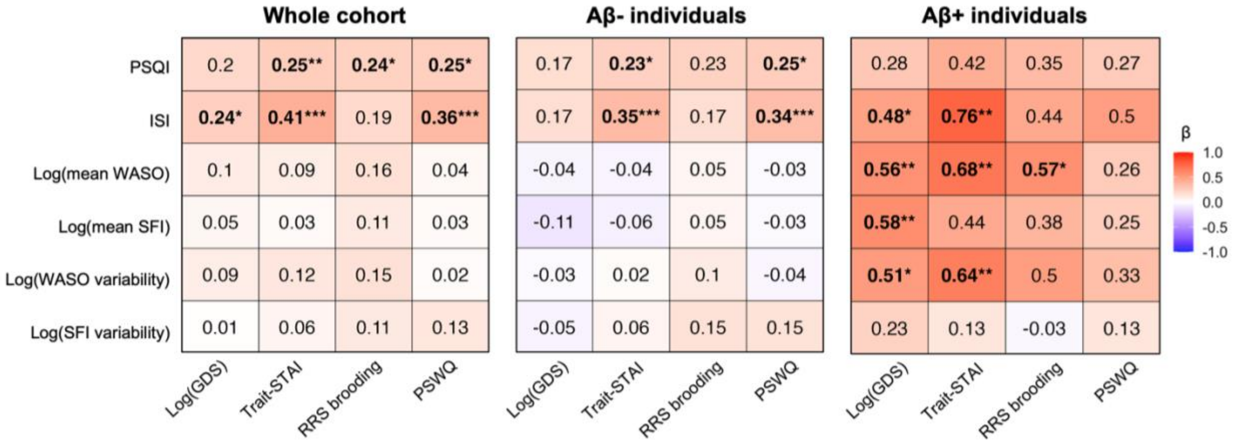


Heatmaps display standardized β coefficients from multiple linear regressions examining associations between self-reported and objective sleep measures (predictors) and psycho-affective symptoms (outcomes), adjusted for age and sex. Analyses were conducted separately in the whole cohort and in subgroups stratified by Aβ status. Results were considered significant when p-values survived an FDR correction for multiple comparisons (*$p_{FDR\text{-}corr}<0.05$, **$p_{FDR\text{-}corr}<0.01$, ***$p_{FDR\text{-}corr}<0.001$). Full statistical details are available in **Table S4**. *Abbreviations: CPAP, continuous positive airway pressure; FDR, false discovery rate; GDS, Geriatric Depression Scale; ISI, Insomnia Severity Index; PSQI, Pittsburgh Sleep Quality Index; PSWQ, Penn State Worry Questionnaire; RRS, Rumination Response Scale; SD, standard deviation; SFI, sleep fragmentation index; STAI, State-Trait Inventory; WASO, wake after sleep onset.*

**<u>Figure S2</u>. Predicted trajectories from significant longitudinal mixed-effects models in Aβ+ individuals, after exclusion of CPAP users.**

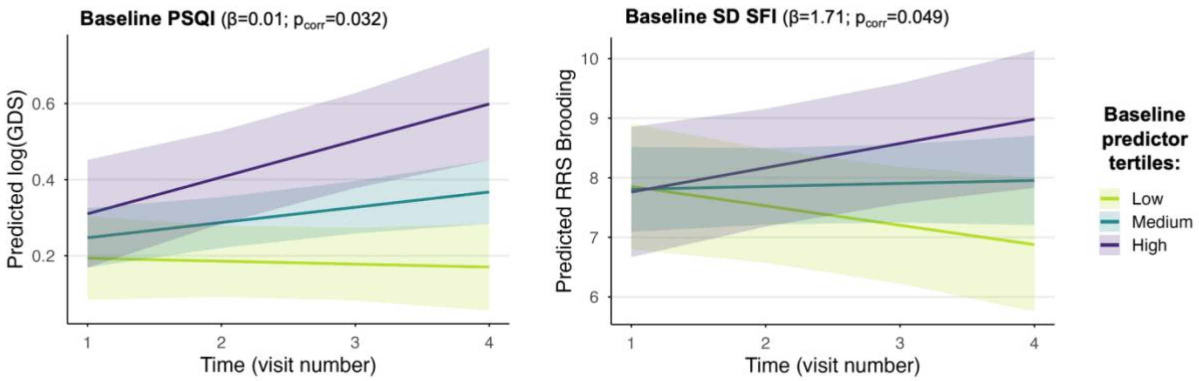


Results of longitudinal mixed-effects models testing the associations between baseline sleep variables and longitudinal changes in psycho-affective scores in Aβ+ individuals. Predicted values represent model-based marginal estimates (± 95% confidence interval) adjusted for age, sex, and the intervention group. Predictions are shown for the mean values of the lower, middle and upper tertiles of each baseline sleep measure. Statistical details are provided in **Table S5**. *Abbreviations: CPAP, continuous positive airway pressure; GDS, Geriatric Depression Scale; ISI, Insomnia Severity Index; PSQI, Pittsburgh Sleep Quality Index; RRS, Rumination Response Scale; SD, standard deviation; SFI, sleep fragmentation index; WASO, wake after sleep onset.*

**Table S4. Cross-sectional associations between sleep measures and psycho-affective symptoms at baseline, after exclusion of CPAP users.**

| Dependent variable | Independent variable | Adjusted $R^2$ | β | 95% CI | SE | t | $P_{unc.}$ | $P_{FDRcorr}$ |
|---|---|---|---|---|---|---|---|---|
| **Whole cohort (n=131)** | | | | | | | | |
| ISI | log_GDS | 0.09 | 0.24 | 0.06–0.41 | 0.09 | 2.71 | 0.008 | **0.046** |
| ISI | Trait-STAI | 0.20 | 0.41 | 0.26–0.57 | 0.08 | 5.22 | <0.001 | **<0.001** |
| ISI | RRS brooding | 0.07 | 0.19 | 0.03–0.36 | 0.09 | 2.28 | 0.024 | 0.073 |
| ISI | PSWQ | 0.17 | 0.36 | 0.20–0.52 | 0.08 | 4.48 | <0.001 | **<0.001** |
| PSQI | log_GDS | 0.09 | 0.20 | 0.03–0.37 | 0.09 | 2.27 | 0.025 | 0.075 |
| PSQI | Trait-STAI | 0.12 | 0.25 | 0.09–0.42 | 0.08 | 3.05 | 0.003 | **0.008** |
| PSQI | RRS brooding | 0.11 | 0.24 | 0.07–0.40 | 0.08 | 2.83 | 0.005 | **0.033** |
| PSQI | PSWQ | 0.12 | 0.25 | 0.08–0.41 | 0.08 | 2.98 | 0.003 | **0.010** |
| log_meanSFI | log_GDS | 0.01 | 0.05 | -0.13–0.23 | 0.09 | 0.54 | 0.593 | 0.711 |
| log_meanSFI | Trait-STAI | 0.01 | 0.03 | -0.14–0.21 | 0.09 | 0.35 | 0.730 | 0.730 |
| log_meanSFI | RRS brooding | 0.02 | 0.11 | -0.07–0.28 | 0.09 | 1.20 | 0.234 | 0.234 |
| log_meanSFI | PSWQ | 0.01 | 0.03 | -0.15–0.21 | 0.09 | 0.33 | 0.741 | 0.836 |
| log_meanWASO | log_GDS | 0.01 | 0.10 | -0.09–0.28 | 0.09 | 1.04 | 0.300 | 0.529 |
| log_meanWASO | Trait-STAI | 0.00 | 0.09 | -0.09–0.26 | 0.09 | 1.00 | 0.318 | 0.477 |
| log_meanWASO | RRS brooding | 0.03 | 0.16 | -0.02–0.33 | 0.09 | 1.80 | 0.074 | 0.143 |
| log_meanWASO | PSWQ | 0.00 | 0.04 | -0.14–0.22 | 0.09 | 0.40 | 0.687 | 0.836 |
| log_SD_SFI | log_GDS | -0.02 | 0.01 | -0.18–0.19 | 0.09 | 0.08 | 0.938 | 0.938 |
| log_SD_SFI | Trait-STAI | -0.01 | 0.06 | -0.12–0.24 | 0.09 | 0.66 | 0.509 | 0.610 |
| log_SD_SFI | RRS brooding | 0.00 | 0.11 | -0.06–0.29 | 0.09 | 1.26 | 0.210 | 0.234 |
| log_SD_SFI | PSWQ | 0.00 | 0.13 | -0.05–0.31 | 0.09 | 1.40 | 0.165 | 0.329 |
| log_SD_WASO | log_GDS | -0.02 | 0.09 | -0.10–0.27 | 0.09 | 0.93 | 0.353 | 0.529 |
| log_SD_WASO | Trait-STAI | -0.01 | 0.12 | -0.05–0.30 | 0.09 | 1.39 | 0.166 | 0.332 |
| log_SD_WASO | RRS brooding | 0.00 | 0.15 | -0.03–0.33 | 0.09 | 1.68 | 0.095 | 0.143 |
| log_SD_WASO | PSWQ | -0.02 | 0.02 | -0.16–0.20 | 0.09 | 0.21 | 0.836 | 0.836 |
| **Aβ- individuals (n=95)** | | | | | | | | |
| ISI | log_GDS | 0.08 | 0.17 | -0.02–0.36 | 0.09 | 1.82 | 0.072 | 0.244 |
| ISI | Trait-STAI | 0.20 | 0.35 | 0.19–0.52 | 0.08 | 4.28 | <0.001 | **<0.001** |
| ISI | RRS brooding | 0.07 | 0.17 | -0.01–0.35 | 0.09 | 1.83 | 0.070 | 0.211 |
| ISI | PSWQ | 0.19 | 0.34 | 0.17–0.51 | 0.08 | 3.99 | <0.001 | **0.001** |
| PSQI | log_GDS | 0.10 | 0.17 | -0.02–0.37 | 0.10 | 1.76 | 0.081 | 0.244 |
| PSQI | Trait-STAI | 0.14 | 0.23 | 0.05–0.41 | 0.09 | 2.59 | 0.011 | **0.033** |
| PSQI | RRS brooding | 0.12 | 0.23 | 0.04–0.42 | 0.10 | 2.44 | 0.017 | 0.101 |
| PSQI | PSWQ | 0.14 | 0.25 | 0.07–0.43 | 0.09 | 2.72 | 0.008 | **0.023** |
| log_meanSFI | log_GDS | 0.01 | -0.11 | -0.31–0.10 | 0.10 | -1.03 | 0.307 | 0.615 |
| log_meanSFI | Trait-STAI | 0.00 | -0.06 | -0.25–0.14 | 0.10 | -0.58 | 0.564 | 0.793 |
| log_meanSFI | RRS brooding | 0.00 | 0.05 | -0.15–0.26 | 0.10 | 0.51 | 0.613 | 0.613 |
| log_meanSFI | PSWQ | 0.00 | -0.03 | -0.23–0.18 | 0.10 | -0.25 | 0.801 | 0.801 |
| log_meanWASO | log_GDS | -0.01 | -0.04 | -0.25–0.17 | 0.11 | -0.40 | 0.693 | 0.802 |

| | | | | | | | | |
|---|---|---|---|---|---|---|---|---|
| log_meanWASO | Trait-STAI | -0.01 | -0.04 | -0.24–0.16 | 0.10 | -0.44 | 0.660 | 0.793 |
| log_meanWASO | RRS brooding | 0.00 | 0.05 | -0.15–0.26 | 0.10 | 0.53 | 0.600 | 0.613 |
| log_meanWASO | PSWQ | -0.01 | -0.03 | -0.23–0.18 | 0.10 | -0.25 | 0.800 | 0.801 |
| log_SD_SFI | log_GDS | -0.03 | -0.05 | -0.25–0.15 | 0.10 | -0.49 | 0.628 | 0.802 |
| log_SD_SFI | Trait-STAI | -0.03 | 0.06 | -0.13–0.25 | 0.09 | 0.62 | 0.534 | 0.793 |
| log_SD_SFI | RRS brooding | 0.00 | 0.15 | -0.04–0.35 | 0.10 | 1.57 | 0.121 | 0.242 |
| log_SD_SFI | PSWQ | 0.00 | 0.15 | -0.05–0.34 | 0.10 | 1.51 | 0.133 | 0.267 |
| log_SD_WASO | log_GDS | -0.03 | -0.03 | -0.23–0.18 | 0.10 | -0.25 | 0.802 | 0.802 |
| log_SD_WASO | Trait-STAI | -0.03 | 0.02 | -0.17–0.22 | 0.10 | 0.22 | 0.824 | 0.824 |
| log_SD_WASO | RRS brooding | -0.02 | 0.10 | -0.10–0.30 | 0.10 | 0.98 | 0.328 | 0.492 |
| log_SD_WASO | PSWQ | -0.03 | -0.04 | -0.24–0.16 | 0.10 | -0.41 | 0.684 | 0.801 |
| **Aβ+ individuals (n=36)** | | | | | | | | |
| ISI | log_GDS | 0.09 | 0.48 | 0.04–0.92 | 0.21 | 2.24 | 0.032 | **0.048** |
| ISI | Trait-STAI | 0.26 | 0.76 | 0.34–1.19 | 0.21 | 3.64 | 0.001 | **0.003** |
| ISI | RRS brooding | 0.06 | 0.44 | -0.02–0.91 | 0.23 | 1.96 | 0.059 | 0.109 |
| ISI | PSWQ | 0.11 | 0.50 | 0.08–0.92 | 0.21 | 2.41 | 0.022 | 0.131 |
| PSQI | log_GDS | 0.00 | 0.28 | -0.12–0.69 | 0.20 | 1.42 | 0.166 | 0.199 |
| PSQI | Trait-STAI | 0.05 | 0.42 | -0.02–0.85 | 0.21 | 1.96 | 0.059 | 0.071 |
| PSQI | RRS brooding | 0.03 | 0.35 | -0.07–0.77 | 0.21 | 1.70 | 0.098 | 0.118 |
| PSQI | PSWQ | 0.00 | 0.27 | -0.13–0.67 | 0.20 | 1.37 | 0.181 | 0.263 |
| log_meanSFI | log_GDS | 0.24 | 0.58 | 0.23–0.93 | 0.17 | 3.36 | 0.002 | **0.007** |
| log_meanSFI | Trait-STAI | 0.10 | 0.44 | 0.03–0.85 | 0.20 | 2.18 | 0.037 | 0.056 |
| log_meanSFI | RRS brooding | 0.06 | 0.38 | -0.04–0.80 | 0.20 | 1.86 | 0.073 | 0.109 |
| log_meanSFI | PSWQ | 0.01 | 0.25 | -0.16–0.66 | 0.20 | 1.25 | 0.219 | 0.263 |
| log_meanWASO | log_GDS | 0.21 | 0.56 | 0.21–0.90 | 0.17 | 3.32 | 0.002 | **0.007** |
| log_meanWASO | Trait-STAI | 0.29 | 0.68 | 0.33–1.04 | 0.17 | 3.95 | <0.001 | **0.002** |
| log_meanWASO | RRS brooding | 0.18 | 0.57 | 0.19–0.94 | 0.18 | 3.09 | 0.004 | **0.025** |
| log_meanWASO | PSWQ | -0.01 | 0.26 | -0.14–0.66 | 0.20 | 1.35 | 0.188 | 0.263 |
| log_SD_SFI | log_GDS | 0.01 | 0.23 | -0.20–0.65 | 0.21 | 1.10 | 0.281 | 0.281 |
| log_SD_SFI | Trait-STAI | -0.02 | 0.13 | -0.33–0.60 | 0.23 | 0.58 | 0.565 | 0.565 |
| log_SD_SFI | RRS brooding | -0.03 | -0.03 | -0.50–0.43 | 0.23 | -0.15 | 0.881 | 0.881 |
| log_SD_SFI | PSWQ | -0.02 | 0.13 | -0.31–0.57 | 0.22 | 0.60 | 0.554 | 0.554 |
| log_SD_WASO | log_GDS | 0.15 | 0.51 | 0.13–0.88 | 0.18 | 2.77 | 0.009 | **0.019** |
| log_SD_WASO | Trait-STAI | 0.22 | 0.64 | 0.26–1.03 | 0.19 | 3.38 | 0.002 | **0.004** |
| log_SD_WASO | RRS brooding | 0.11 | 0.50 | 0.09–0.91 | 0.20 | 2.48 | 0.019 | 0.056 |
| log_SD_WASO | PSWQ | 0.02 | 0.33 | -0.08–0.75 | 0.20 | 1.63 | 0.114 | 0.263 |

Results from multiple regression analyses examining cross-sectional associations between self-reported or objective sleep measures and psycho-affective symptoms at baseline, after excluding CPAP users. Data are reported as standardized β coefficients with corresponding 95% confidence intervals (CI), and both uncorrected ($P_{unc.}$) and FDR-corrected ($P_{FDRcorr}$) p-values. Results surviving an FDR correction ($P_{FDRcorr}<0.05$) are indicated in bold. All models were adjusted for age and sex, and analyses were performed in the whole cohort and in subgroups stratified by Aβ status. *Abbreviations: Aβ, amyloid; CPAP, continuous positive airway pressure; FDR, false discovery rate; GDS, Geriatric Depression Scale; ISI, Insomnia*

*Severity Index; PSQI, Pittsburgh Sleep Quality Index; PSWQ, Penn State Worry Questionnaire; RRS, Ruminative Response Scale; SD, standard deviation; SFI, sleep fragmentation index; STAI, State-Trait Anxiety Inventory; WASO, wake after sleep onset.*

**Table S5. Associations between baseline sleep measures and longitudinal psycho-affective changes, after exclusion of CPAP users.**

| Outcome | Time × Predictor interaction | Estimate | SE | t | $P_{unc.}$ | $P_{FDRcorr}$ |
|---|---|---|---|---|---|---|
| **WHOLE COHORT (n=121)** | | | | | | |
| log_GDS | Time x baseline_ISI | 0.001 | 0.002 | 0.55 | 0.581 | 0.833 |
| log_GDS | Time x baseline_PSQI | 0.001 | 0.003 | 0.49 | 0.628 | 0.833 |
| log_GDS | Time x baseline_log_meanWASO | 0.02 | 0.06 | 0.39 | 0.694 | 0.833 |
| log_GDS | Time x baseline_log_meanSFI | 0.09 | 0.06 | 1.45 | 0.148 | 0.833 |
| log_GDS | Time x baseline_log_SD_WASO | 0.01 | 0.04 | 0.17 | 0.869 | 0.869 |
| log_GDS | Time x baseline_log_SD_SFI | 0.06 | 0.05 | 1.04 | 0.299 | 0.833 |
| PSWQ | Time x baseline_ISI | -0.06 | 0.04 | -1.42 | 0.156 | 0.469 |
| PSWQ | Time x baseline_PSQI | -0.10 | 0.07 | -1.49 | 0.137 | 0.469 |
| PSWQ | Time x baseline_log_meanWASO | -1.14 | 1.32 | -0.86 | 0.392 | 0.593 |
| PSWQ | Time x baseline_log_meanSFI | -1.28 | 1.51 | -0.85 | 0.395 | 0.593 |
| PSWQ | Time x baseline_log_SD_WASO | -0.44 | 0.99 | -0.44 | 0.661 | 0.793 |
| PSWQ | Time x baseline_log_SD_SFI | -0.24 | 1.29 | -0.19 | 0.852 | 0.852 |
| RRS brooding | Time x baseline_ISI | 0.02 | 0.01 | 1.56 | 0.119 | 0.211 |
| RRS brooding | Time x baseline_PSQI | 0.01 | 0.02 | 0.48 | 0.634 | 0.634 |
| RRS brooding | Time x baseline_log_meanWASO | 0.24 | 0.39 | 0.61 | 0.545 | 0.634 |
| RRS brooding | Time x baseline_log_meanSFI | 0.65 | 0.44 | 1.48 | 0.141 | 0.211 |
| RRS brooding | Time x baseline_log_SD_WASO | 0.47 | 0.29 | 1.61 | 0.109 | 0.211 |
| RRS brooding | Time x baseline_log_SD_SFI | 0.79 | 0.38 | 2.11 | 0.036 | 0.211 |
| Trait-STAI | Time x baseline_ISI | -0.03 | 0.03 | -0.89 | 0.373 | 0.627 |
| Trait-STAI | Time x baseline_PSQI | 0.01 | 0.05 | 0.13 | 0.894 | 0.894 |
| Trait-STAI | Time x baseline_log_meanWASO | -0.55 | 0.95 | -0.58 | 0.564 | 0.677 |
| Trait-STAI | Time x baseline_log_meanSFI | 1.10 | 1.08 | 1.02 | 0.310 | 0.627 |
| Trait-STAI | Time x baseline_log_SD_WASO | 0.58 | 0.71 | 0.81 | 0.418 | 0.627 |
| Trait-STAI | Time x baseline_log_SD_SFI | 2.28 | 0.92 | 2.48 | 0.014 | 0.083 |
| **Aβ- INDIVIDUALS (n=89)** | | | | | | |
| log_GDS | Time x baseline_ISI | -0.001 | 0.002 | -0.46 | 0.646 | 0.775 |
| log_GDS | Time x baseline_PSQI | -0.004 | 0.003 | -1.05 | 0.293 | 0.664 |
| log_GDS | Time x baseline_log_meanWASO | 0.06 | 0.07 | 0.97 | 0.332 | 0.664 |
| log_GDS | Time x baseline_log_meanSFI | 0.17 | 0.07 | 2.26 | 0.025 | 0.149 |
| log_GDS | Time x baseline_log_SD_WASO | 0.004 | 0.05 | 0.09 | 0.931 | 0.931 |
| log_GDS | Time x baseline_log_SD_SFI | 0.04 | 0.07 | 0.60 | 0.548 | 0.775 |
| PSWQ | Time x baseline_ISI | -0.09 | 0.05 | -1.58 | 0.115 | 0.344 |
| PSWQ | Time x baseline_PSQI | -0.20 | 0.08 | -2.45 | 0.015 | 0.091 |
| PSWQ | Time x baseline_log_meanWASO | -0.78 | 1.56 | -0.50 | 0.615 | 0.815 |
| PSWQ | Time x baseline_log_meanSFI | -1.54 | 1.78 | -0.87 | 0.388 | 0.775 |
| PSWQ | Time x baseline_log_SD_WASO | 0.21 | 1.21 | 0.17 | 0.865 | 0.865 |
| PSWQ | Time x baseline_log_SD_SFI | 0.68 | 1.63 | 0.41 | 0.679 | 0.815 |

| | | | | | | |
|---|---|---|---|---|---|---|
| RRS brooding | Time x baseline_ISI | 0.01 | 0.02 | 0.38 | 0.706 | 0.714 |
| RRS brooding | Time x baseline_PSQI | -0.02 | 0.02 | -0.64 | 0.521 | 0.714 |
| RRS brooding | Time x baseline_log_meanWASO | 0.17 | 0.46 | 0.37 | 0.714 | 0.714 |
| RRS brooding | Time x baseline_log_meanSFI | 0.41 | 0.52 | 0.80 | 0.424 | 0.714 |
| RRS brooding | Time x baseline_log_SD_WASO | 0.15 | 0.35 | 0.42 | 0.672 | 0.714 |
| RRS brooding | Time x baseline_log_SD_SFI | 0.34 | 0.47 | 0.71 | 0.480 | 0.714 |
| Trait-STAI | Time x baseline_ISI | 0.001 | 0.03 | 0.03 | 0.980 | 0.992 |
| Trait-STAI | Time x baseline_PSQI | 0.03 | 0.05 | 0.49 | 0.628 | 0.942 |
| Trait-STAI | Time x baseline_log_meanWASO | 0.01 | 1.00 | 0.01 | 0.992 | 0.992 |
| Trait-STAI | Time x baseline_log_meanSFI | 1.27 | 1.14 | 1.12 | 0.263 | 0.673 |
| Trait-STAI | Time x baseline_log_SD_WASO | 0.74 | 0.77 | 0.96 | 0.336 | 0.673 |
| Trait-STAI | Time x baseline_log_SD_SFI | 2.22 | 1.04 | 2.14 | 0.033 | 0.200 |
| **Aβ+ INDIVIDUALS (n=32)** | | | | | | |
| log_GDS | Time x baseline_ISI | 0.005 | 0.003 | 1.72 | 0.089 | 0.268 |
| log_GDS | Time x baseline_PSQI | 0.01 | 0.004 | 2.87 | 0.005 | **0.032** |
| log_GDS | Time x baseline_log_meanWASO | -0.10 | 0.10 | -1.00 | 0.319 | 0.383 |
| log_GDS | Time x baseline_log_meanSFI | -0.14 | 0.11 | -1.19 | 0.238 | 0.383 |
| log_GDS | Time x baseline_log_SD_WASO | 0.02 | 0.07 | 0.22 | 0.825 | 0.825 |
| log_GDS | Time x baseline_log_SD_SFI | 0.09 | 0.09 | 1.03 | 0.305 | 0.383 |
| PSWQ | Time x baseline_ISI | -0.02 | 0.07 | -0.22 | 0.829 | 0.829 |
| PSWQ | Time x baseline_PSQI | 0.13 | 0.12 | 1.08 | 0.285 | 0.606 |
| PSWQ | Time x baseline_log_meanWASO | -2.17 | 2.53 | -0.86 | 0.393 | 0.606 |
| PSWQ | Time x baseline_log_meanSFI | -0.64 | 2.88 | -0.22 | 0.826 | 0.829 |
| PSWQ | Time x baseline_log_SD_WASO | -1.87 | 1.83 | -1.02 | 0.309 | 0.606 |
| PSWQ | Time x baseline_log_SD_SFI | -1.85 | 2.21 | -0.84 | 0.404 | 0.606 |
| RRS brooding | Time x baseline_ISI | 0.04 | 0.02 | 2.14 | 0.035 | 0.071 |
| RRS brooding | Time x baseline_PSQI | 0.06 | 0.03 | 1.85 | 0.068 | 0.102 |
| RRS brooding | Time x baseline_log_meanWASO | 0.41 | 0.75 | 0.55 | 0.587 | 0.587 |
| RRS brooding | Time x baseline_log_meanSFI | 1.36 | 0.84 | 1.62 | 0.109 | 0.131 |
| RRS brooding | Time x baseline_log_SD_WASO | 1.25 | 0.52 | 2.38 | 0.020 | 0.059 |
| RRS brooding | Time x baseline_log_SD_SFI | 1.71 | 0.63 | 2.71 | 0.008 | **0.049** |
| Trait-STAI | Time x baseline_ISI | -0.08 | 0.06 | -1.36 | 0.176 | 0.699 |
| Trait-STAI | Time x baseline_PSQI | -0.05 | 0.11 | -0.47 | 0.641 | 0.806 |
| Trait-STAI | Time x baseline_log_meanWASO | -2.33 | 2.33 | -1.00 | 0.319 | 0.699 |
| Trait-STAI | Time x baseline_log_meanSFI | 0.71 | 2.66 | 0.27 | 0.791 | 0.806 |
| Trait-STAI | Time x baseline_log_SD_WASO | -0.42 | 1.69 | -0.25 | 0.806 | 0.806 |
| Trait-STAI | Time x baseline_log_SD_SFI | 1.91 | 2.03 | 0.94 | 0.349 | 0.699 |

Results of longitudinal mixed-effects models testing whether baseline sleep quality predicted longitudinal changes in psycho-affective symptoms over time, after excluding CPAP users. Data are reported as estimated standardized β coefficients (estimates), standard error (SE), t-value, and both uncorrected ($P_{unc.}$) and FDR-corrected ($P_{FDRcorr}$) p-values. Results surviving FDR correction ($P_{FDRcorr}<0.05$) are indicated in bold. All models were adjusted for age, sex, and the intervention group. Analyses were performed in the whole cohort and in subgroups stratified by Aβ status. *Abbreviations: Aβ, amyloid; CPAP, continuous positive airway pressure; FDR, false discovery rate; GDS, Geriatric Depression Scale; ISI, Insomnia Severity Index; PSQI, Pittsburgh Sleep Quality Index; PSWQ, Penn State Worry Questionnaire; RRS, Ruminative Response Scale; SD, standard deviation; SFI, sleep fragmentation index; STAI, State-Trait Anxiety Inventory; WASO, wake after sleep onset.*

**Table S6. Associations between baseline psycho-affective symptoms and sleep changes over time, after exclusion of CPAP users.**

| Outcome | Time × Predictor interaction | Estimate | SE | t | $P_{unc.}$ | $P_{FDRcorr}$ |
|---|---|---|---|---|---|---|
| **WHOLE COHORT (n=121)** | | | | | | |
| ISI | Time x baseline_logGDS | 0.66 | 0.48 | 1.38 | 0.169 | 0.544 |
| ISI | Time x baseline_Trait-STAI | 0.007 | 0.02 | 0.44 | 0.662 | 0.662 |
| ISI | Time x baseline_PSWQ | -0.01 | 0.01 | -1.10 | 0.272 | 0.544 |
| ISI | Time x baseline_RRS brooding | 0.04 | 0.05 | 0.69 | 0.491 | 0.655 |
| PSQI | Time x baseline_logGDS | 0.58 | 0.34 | 1.72 | 0.087 | 0.348 |
| PSQI | Time x baseline_Trait-STAI | 0.01 | 0.01 | 1.04 | 0.301 | 0.601 |
| PSQI | Time x baseline_PSWQ | 0.001 | 0.01 | 0.09 | 0.928 | 0.928 |
| PSQI | Time x baseline_RRS brooding | 0.01 | 0.04 | 0.28 | 0.778 | 0.928 |
| log_meanSFI | Time x baseline_logGDS | 0.007 | 0.02 | 0.46 | 0.646 | 0.821 |
| log_meanSFI | Time x baseline_Trait-STAI | 3.07E-04 | 0.001 | 0.57 | 0.571 | 0.821 |
| log_meanSFI | Time x baseline_PSWQ | -8.38E-05 | 3.50E-04 | -0.24 | 0.811 | 0.821 |
| log_meanSFI | Time x baseline_RRS brooding | -3.93E-04 | 0.002 | -0.23 | 0.821 | 0.821 |
| log_meanWASO | Time x baseline_logGDS | 0.004 | 0.02 | 0.25 | 0.800 | 0.800 |
| log_meanWASO | Time x baseline_Trait-STAI | -3.38E-04 | 0.001 | -0.62 | 0.536 | 0.800 |
| log_meanWASO | Time x baseline_PSWQ | -1.68E-04 | 3.53E-04 | -0.48 | 0.634 | 0.800 |
| log_meanWASO | Time x baseline_RRS brooding | -0.002 | 0.002 | -1.05 | 0.295 | 0.800 |
| log_SD_SFI | Time x baseline_logGDS | 0.01 | 0.03 | 0.39 | 0.701 | 0.701 |
| log_SD_SFI | Time x baseline_Trait-STAI | 0.001 | 0.001 | 1.54 | 0.124 | 0.248 |
| log_SD_SFI | Time x baseline_PSWQ | 4.94E-04 | 0.001 | 0.86 | 0.389 | 0.518 |
| log_SD_SFI | Time x baseline_RRS brooding | 0.005 | 0.003 | 1.94 | 0.054 | 0.216 |
| log_SD_WASO | Time x baseline_logGDS | 0.003 | 0.03 | 0.09 | 0.930 | 0.930 |
| log_SD_WASO | Time x baseline_Trait-STAI | -0.001 | 0.001 | -1.35 | 0.177 | 0.364 |
| log_SD_WASO | Time x baseline_PSWQ | -0.001 | 0.001 | -0.93 | 0.352 | 0.470 |
| log_SD_WASO | Time x baseline_RRS brooding | -0.004 | 0.003 | -1.34 | 0.182 | 0.364 |
| **Aβ- INDIVIDUALS (n=89)** | | | | | | |
| ISI | Time x baseline_logGDS | 0.65 | 0.57 | 1.14 | 0.254 | 0.508 |
| ISI | Time x baseline_Trait-STAI | 0.01 | 0.02 | 0.58 | 0.562 | 0.562 |
| ISI | Time x baseline_PSWQ | -0.02 | 0.01 | -1.36 | 0.177 | 0.508 |
| ISI | Time x baseline_RRS brooding | 0.04 | 0.06 | 0.66 | 0.509 | 0.562 |
| PSQI | Time x baseline_logGDS | 0.37 | 0.36 | 1.02 | 0.311 | 0.758 |
| PSQI | Time x baseline_Trait-STAI | 0.01 | 0.01 | 0.88 | 0.379 | 0.758 |
| PSQI | Time x baseline_PSWQ | -0.004 | 0.01 | -0.54 | 0.590 | 0.787 |
| PSQI | Time x baseline_RRS brooding | 0.005 | 0.04 | 0.11 | 0.910 | 0.910 |
| log_meanSFI | Time x baseline_logGDS | 0.02 | 0.02 | 0.96 | 0.336 | 0.855 |
| log_meanSFI | Time x baseline_Trait-STAI | 0.001 | 0.001 | 0.80 | 0.427 | 0.855 |
| log_meanSFI | Time x baseline_PSWQ | -9.65E-06 | 4.13E-04 | -0.02 | 0.981 | 0.981 |
| log_meanSFI | Time x baseline_RRS brooding | 0.001 | 0.002 | 0.24 | 0.807 | 0.981 |

| | | | | | | |
|---|---|---|---|---|---|---|
| log_meanWASO | Time x baseline_logGDS | 0.01 | 0.02 | 0.70 | 0.486 | 0.869 |
| log_meanWASO | Time x baseline_Trait-STAI | -1.87E-04 | 0.001 | -0.30 | 0.764 | 0.869 |
| log_meanWASO | Time x baseline_PSWQ | 6.57E-05 | 3.99E-04 | 0.16 | 0.869 | 0.869 |
| log_meanWASO | Time x baseline_RRS brooding | -4.06E-04 | 0.002 | -0.20 | 0.838 | 0.869 |
| log_SD_SFI | Time x baseline_logGDS | 0.04 | 0.03 | 1.37 | 0.174 | 0.231 |
| log_SD_SFI | Time x baseline_Trait-STAI | 0.002 | 0.001 | 2.18 | 0.031 | 0.091 |
| log_SD_SFI | Time x baseline_PSWQ | 0.001 | 0.001 | 0.82 | 0.416 | 0.416 |
| log_SD_SFI | Time x baseline_RRS brooding | 0.01 | 0.003 | 2.02 | 0.045 | 0.091 |
| log_SD_WASO | Time x baseline_logGDS | 0.03 | 0.03 | 1.01 | 0.315 | 0.671 |
| log_SD_WASO | Time x baseline_Trait-STAI | -0.001 | 0.001 | -0.67 | 0.503 | 0.671 |
| log_SD_WASO | Time x baseline_PSWQ | -1.32E-04 | 0.001 | -0.19 | 0.853 | 0.853 |
| log_SD_WASO | Time x baseline_RRS brooding | -0.003 | 0.004 | -0.75 | 0.456 | 0.671 |
| **Aβ+ INDIVIDUALS (n=32)** | | | | | | |
| ISI | Time x baseline_logGDS | 0.41 | 0.84 | 0.49 | 0.624 | 0.906 |
| ISI | Time x baseline_Trait-STAI | -0.03 | 0.03 | -0.94 | 0.351 | 0.906 |
| ISI | Time x baseline_PSWQ | 0.003 | 0.02 | 0.17 | 0.866 | 0.906 |
| ISI | Time x baseline_RRS brooding | 0.01 | 0.09 | 0.12 | 0.906 | 0.906 |
| PSQI | Time x baseline_logGDS | 1.05 | 0.78 | 1.35 | 0.182 | 0.729 |
| PSQI | Time x baseline_Trait-STAI | 0.001 | 0.03 | 0.05 | 0.961 | 0.961 |
| PSQI | Time x baseline_PSWQ | 0.02 | 0.02 | 0.89 | 0.380 | 0.759 |
| PSQI | Time x baseline_RRS brooding | 0.02 | 0.08 | 0.28 | 0.778 | 0.961 |
| log_meanSFI | Time x baseline_logGDS | -0.03 | 0.03 | -0.94 | 0.351 | 0.702 |
| log_meanSFI | Time x baseline_Trait-STAI | -2.39E-04 | 0.001 | -0.24 | 0.812 | 0.812 |
| log_meanSFI | Time x baseline_PSWQ | -3.14E-04 | 0.001 | -0.48 | 0.636 | 0.812 |
| log_meanSFI | Time x baseline_RRS brooding | -0.003 | 0.003 | -1.03 | 0.307 | 0.702 |
| log_meanWASO | Time x baseline_logGDS | -0.03 | 0.03 | -0.78 | 0.436 | 0.436 |
| log_meanWASO | Time x baseline_Trait-STAI | -0.001 | 0.001 | -0.85 | 0.397 | 0.436 |
| log_meanWASO | Time x baseline_PSWQ | -0.001 | 0.001 | -1.47 | 0.147 | 0.294 |
| log_meanWASO | Time x baseline_RRS brooding | -0.007 | 0.004 | -1.87 | 0.067 | 0.269 |
| log_SD_SFI | Time x baseline_logGDS | -0.10 | 0.05 | -1.84 | 0.072 | 0.289 |
| log_SD_SFI | Time x baseline_Trait-STAI | -0.002 | 0.002 | -0.99 | 0.328 | 0.656 |
| log_SD_SFI | Time x baseline_PSWQ | 1.13E-04 | 0.001 | 0.09 | 0.931 | 0.931 |
| log_SD_SFI | Time x baseline_RRS brooding | 0.002 | 0.01 | 0.27 | 0.785 | 0.931 |
| log_SD_WASO | Time x baseline_logGDS | -0.11 | 0.07 | -1.57 | 0.122 | 0.151 |
| log_SD_WASO | Time x baseline_Trait-STAI | -0.005 | 0.003 | -1.88 | 0.066 | 0.151 |
| log_SD_WASO | Time x baseline_PSWQ | -0.003 | 0.002 | -1.77 | 0.083 | 0.151 |
| log_SD_WASO | Time x baseline_RRS brooding | -0.01 | 0.01 | -1.46 | 0.151 | 0.151 |

Results of longitudinal mixed-effects models testing whether baseline psycho-affective symptoms predicted longitudinal changes in sleep quality over time, after excluding CPAP users. Data are reported as estimated standardized β coefficients (estimates), standard error (SE), t-value, and both uncorrected ($P_{unc.}$) and FDR-corrected ($P_{FDRcorr}$) p-values. Results surviving FDR correction ($P_{FDRcorr}$<0.05) are indicated in bold. All models were adjusted for age, sex, and the intervention group. Analyses were performed in the whole cohort and in subgroups stratified by Aβ status. *Abbreviations: Aβ, amyloid; CPAP, continuous positive airway pressure; FDR, false discovery rate; GDS, Geriatric Depression Scale; ISI, Insomnia Severity Index; PSQI, Pittsburgh Sleep Quality Index; PSWQ, Penn State Worry Questionnaire; RRS, Ruminative Response Scale; SD, standard deviation; SFI, sleep fragmentation index; STAI, State-Trait Anxiety Inventory; WASO, wake after sleep onset.*

**<u>Appendix</u>. The Medit-Ageing Research Group.**

| Name, Degree | Location | Role | Contribution |
|---|---|---|---|
| Florence Allais, BA | EUCLID/F-CRIN Clinical Trials Platform, Bordeaux, France | Data manager | Data management |
| Eider M. Arenaza-Urquijo, PhD | Institut National de la Santé et de la Recherche Médicale, Caen, France | Postdoctoral researcher | Study design, acquisition, analysis, and/or interpretation of data |
| Julien Asselineau, PhD | EUCLID/F-CRIN Clinical Trials Platform, Bordeaux, France | Statistician | Statistical analysis of data |
| Sebastian Baez Lugo, MSc | University of Geneva, Geneva, Switzerland | PhD student | Acquisition, analysis, or interpretation of data |
| Alexandre Bejanin, PhD | Institut National de la Santé et de la Recherche, Médicale, Caen, France | Postdoctoral researcher | Acquisition, analysis, and/or interpretation of data |
| Anne Chocat, MD | Institut National de la Santé et de la Recherche, Médicale, Caen, France | Physician | Supervision of participants recruitment |
| Fabienne Collette, PhD | University of Liege, Liege, Belgium | Group leader | Obtained funding and study design |
| Marion Delarue, MSc | Institut National de la Santé et de la Recherche, Médicale, Caen, France | Neuropsychologist | Acquisition, analysis, and/or interpretation of data |
| Hélène Esperou, MD | Institut National de la Santé et de la Recherche, Médicale, Paris, France | Group Leader | Sponsor – monitoring of the trial |
| Eglantine Ferrand-Devouge, MD | Institut National de la Santé et de la Recherche, Médicale, Caen, France | Physician | Supervision of participants recruitment |
| Robin de Flores, PhD | Institut National de la Santé et de la Recherche Médicale, Caen, France | Postdoctoral researcher | Acquisition, analysis, and/or interpretation of data |
| Eric Frison, MD, PhD | EUCLID/F-CRIN Clinical Trials Platform, Bordeaux, France | Methodologist | Study design and interpretation of data |
| Julie Gonneaud, PhD | Institut National de la Santé et de la Recherche, Médicale, Caen, France | Postdoctoral researcher | Acquisition, analysis, and/or interpretation of data |
| Olga Klimecki, PhD | University of Geneva, Geneva, Switzerland | Group leader | Obtained funding and study design |

| | | | |
|---|---|---|---|
| Brigitte Landeau, MSc | Institut National de la Santé et de la Recherche, Médicale, Caen, France | Neuroimaging developer engineer | Neuroimaging data processing |
| Gwendoline Le Du, MSc | Institut National de la Santé et de la Recherche, Médicale, Caen, France | Clinical research assistant | Acquisition, analysis, and/or interpretation of data, and administrative, technical, and/or material support |
| Valérie Lefranc, BA | Institut National de la Santé et de la Recherche, Médicale, Caen, France | Clinical research assistant | Acquisition, analysis, and/or interpretation of data, and administrative, technical, and/or material support |
| Antoine Lutz, PhD | Institut National de la Santé et de la Recherche Médicale, Lyon, France | Group leader | Obtained funding and study design |
| Florence Mézenge, BA | Institut National de la Santé et de la Recherche, Médicale, Caen, France | Clinical research assistant | Acquisition, analysis, and/or interpretation of data, and administrative, technical, and/or material support |
| Jose-Luis Molinuevo, MD, PhD | Institut d'Investigacions Biomèdiques August Pi i Sunyer, Barcelona, Spain | Group leader | Obtained funding and study design |
| Inès Moulinet, MSc | Institut National de la Santé et de la Recherche, Médicale, Caen, France | PhD student | Acquisition, analysis, and/or interpretation of data |
| Léo Paly, MSc | Institut National de la Santé et de la Recherche, Médicale, Caen, France | Neuropsychologist | Acquisition, analysis, and/or interpretation of data |
| Géraldine Poisnel, PhD | Institut National de la Santé et de la Recherche Médicale, Caen, France | Research engineer /Researcher | Obtained funding and study design, acquisition, analysis, and/or interpretation of data |
| Anne Quillard, MD | Institut National de la Santé et de la Recherche, Médicale, Caen, France | Physician | Supervision of participants recruitment |
| Florence Requier, MSc | University of Liege, Liege, Belgium | PhD student | Acquisition, analysis, and/or interpretation of data |
| Eric Salmon, MD, PhD | University of Liege, Liege, Belgium | Group leader | Obtained funding and study design |

| | | | |
|---|---|---|---|
| Siya Sherif, PhD | Institut National de la Santé et de la Recherche, Médicale, Caen, France | Research engineer | Acquisition, analysis, and/or interpretation of data and administrative, technical, and/or material support |
| Matthieu Vanhoutte, PhD | Institut National de la Santé et de la Recherche, Médicale, Caen, France | Postdoctoral researcher | Acquisition, analysis, and/or interpretation of data |
| Patrik Vuillemier, MD | University of Geneva, Geneva, Switzerland | Group leader | Obtained funding and study design |
| Miranka Wirth, PhD | Deutsches Zentrum für Neurodegenerative Erkrankungen, Dresden, Germany | Group leader | Obtained funding and study design |
